# Generative AI Mitigates Representation Bias and Improves Model Fairness Through Synthetic Health Data

**DOI:** 10.1101/2023.09.26.23296163

**Authors:** Raffaele Marchesi, Nicolo Micheletti, Nicholas I-Hsien Kuo, Sebastiano Barbieri, Giuseppe Jurman, Venet Osmani

**Affiliations:** Fondazione Bruno Kessler Research Institute, Italy; Department of Mathematics, University of Pavia, Italy; Department of Computer Science, University of Manchester, United Kingdom; Centre for Big Data Research in Health, University of New South Wales, Australia; Queensland Digital Health Centre, University of Queensland, Australia; Digital Environment Research Institute (DERI), Queen Mary University of London, United Kingdom

**Keywords:** generative adversarial networks, GAN, synthetic data, resampling, oversampling, data bias, underrepresentation, health data, algorithmic fairness, ethnicity, gender

## Abstract

Representation bias in health data can lead to unfair decisions and compromise the generalisability of research findings. As a consequence, underrepresented subpopulations, such as those from specific ethnic backgrounds or genders, do not benefit equally from clinical discoveries. Several approaches have been developed to mitigate representation bias, ranging from simple resampling methods, such as SMOTE, to recent approaches based on generative adversarial networks (GAN). However, generating high-dimensional time-series synthetic health data remains a significant challenge. In response, we devised a novel architecture (CA-GAN) that synthesises authentic, high-dimensional time series data. CA-GAN outperforms state-of-the-art methods in a qualitative and a quantitative evaluation while avoiding mode collapse, a serious GAN failure. We perform evaluation using 7535 patients with hypotension and sepsis from two diverse, real-world clinical datasets. We show that synthetic data generated by our CA-GAN improves model fairness in Black patients as well as female patients when evaluated separately for each subpopulation. Furthermore, CA-GAN generates authentic data of the minority class while faithfully maintaining the original distribution of data, resulting in improved performance in a downstream predictive task.

**Author summary:** Doctors and other healthcare professionals are increasingly using Artificial Intelligence (AI) to make better decisions about patients’ diagnosis, suggest optimal treatments, and estimate patients’ future health risks. These AI systems learn from existing health data which might not accurately reflect the health of everyone, particularly people from certain racial or ethnic groups, genders, or those with lower incomes. This can mean the AI doesn’t work as well for these groups and could even make existing health disparities worse. To address this, we have developed a purposely built AI software that can create *synthetic* patient data. Synthetic data created by our software mimics real patient data without actually copying them, protecting patients’ privacy. Using our synthetic data results in more representative dataset of all groups, and ensures that AI algorithms learn to be fairer for all patients.

## 1 Introduction

Clinical practice is poised to benefit from developments in machine learning as data-driven digital health technologies transform health care [1]. Digital health can catalyse the World Health Organisation’s (WHO) vision of promoting equitable, affordable, and universal access to health and care [2]. However, as machine learning methods increasingly weave themselves into the societal fabric, critical issues related to fairness and algorithmic bias in decision-making are coming to light. Algorithmic bias can originate from diverse sources, including socio-economic factors, where income disparities between ethno-racial groups are reflected in algorithms deciding which patients need care [3]. Bias can also originate from the underrepresentation of particular demographics, such as ethnicity, gender, and age in the datasets used to develop machine learning models, known as health data poverty [4]. Health data poverty impedes underrepresented subpopulations from benefiting from clinical discoveries, compromising the generalisability of research findings and leading to representation bias that can compound health disparities.

Machine learning community has developed several approaches to mitigate representation bias, with data resampling being the most widely used. Over-sampling generates representative synthetic data from the underrepresented subpopulation (minority class), resulting in a similar or equal representation. Synthetic Minority Over-sampling TEchnique (SMOTE) [5] is a representative example of this method, where synthetic samples lie between a randomly selected data sample and its randomly selected neighbour (using k-nearest neighbour algorithm [6]). SMOTE and related methods [7–9] are popular approaches due to their simplicity and computational efficiency.

However, SMOTE, when used with high-dimensional time-series data, may decrease data variability and introduce correlation between samples [10–12]. In response, alternative approaches based on Generative Adversarial Networks (GAN) are gaining ground [13–17]. GANs have shown incredible results in generating realistic images [18], text [19], and speech [20] in addition to improving privacy [21]. While GANs address some of the issues of SMOTE-based approaches, the generation of high-dimensional time-series data remains a significant research challenge [22–24].

To address this challenge, we propose a new generative architecture called Conditional Augmentation GAN (CA-GAN). Our CA-GAN extends Wasserstein GAN with Gradient Penalty [25, 26], presented in the Health Gym study [27] (referred to in this paper as WGAN-GP*). However, our work has a different objective. Instead of generating new synthetic datasets, we condition our GAN to augment the minority class only, while maintaining correlations between the variables and correlations over time, in contrast to the recent work [28]. As a result, CA-GAN captures the distribution of the overall dataset, including the majority class. We compare the performance of our CA-GAN with WGAN-GP* and SMOTE in generating synthetic data of patients of an under-represented ethnicity (Black patients in our case) as well as gender (female). We use two critical care datasets comprising acute hypotension (n=3343) and sepsis (n=4192), resulting in 7535 patients overall.

Our datasets include both categorical and continuous variables with diverse distributions and are derived from the well-studied MIMIC-III critical care database [29]. These datasets were chosen to allow direct comparison with state of the art approaches, our architecture is data agnostic.

Our work makes the following contributions: (1) We propose a new CA-GAN architecture that addresses the shortcomings of traditional and the state-of-the-art methods in generating high-dimensional, time-series, synthetic data, using two real-world datasets. (2) Our multi-metric evaluation using qualitative and quantitative methods demonstrate superior performance of CA-GAN with respect to the state of the art architecture, while avoiding mode-collapse, a significant GAN failure. (3) We evaluate our CA-GAN against SMOTE, a naive but cost-effective resampling method, however limited in the synthesis of authentic data. (4) We show the impact of synthetic data in improving model performance for the minority (underrepresented) class, resulting in a fairer model between Black and White ethnicities. (5) We also show that CA-GAN can synthesise realistic clinical data of specific ethnicity and gender, improving the performance in a downstream predictive task.

## 2 Results

We primarily focus on multi-metric evaluation of synthetic data generated by our CA-GAN architecture in comparison to the data generated by state-of-the-art WGAN-GP* architecture and the popular SMOTE approach. We provide a separate analysis on the impact of synthetic data generated by our architecture to mitigate representation bias and improve model fairness in Section 2.5. Considering significant challenges in evaluating generative models in general, [30], and high-dimensional time-series data in particular [23], we adopted a holistic approach to evaluating our work based on both qualitative and quantitative methods. We present the results of the data generated comparing the performance of the three methods in augmenting the underrepresented (minority) class, namely Black ethnicity and female gender.

### 2.1 Qualitative evaluation

To gain initial insights into the obtained results, we conduct a qualitative evaluation employing visual representation methods that show the coverage of synthetic data with respect to the real data. We use Principal Component Analysis (PCA) to project the real and synthetic data onto a two-dimensional space. We also use t-distributed Stochastic Neighbor Embedding (t-SNE) [31] to plot both real and synthetic datasets in a two-dimensional latent space while preserving the local neighbourhood relationships between data points. To compare the performance between the methods and ensure a consistent visualisation of the real data, we have computed a common t-SNE embedding. In Appendix G we present the results of Uniform Manifold Approximation and Projection (UMAP) [32], which offers better preservation of the global structure of the dataset when compared to t-SNE. The parameters of t-SNE and UMAP are the same for all three methods as shown in Appendix C.

The results are illustrated in Figure 1 (acute hypotension) and Figure 2 (sepsis). Synthetic data generated by CA-GAN exhibits significant overlap with the real data, indicating our model’s ability to accurately capture the underlying structure of real data. This is especially evident in PCA, where the representations reveal that the synthetic data generated by CA-GAN provides the best overall coverage of the real data distribution. Further evidence is provided from the marginal distributions. For the acute hypotension dataset (Figure 1), both WGAN-GP* and SMOTE show evidence of mode collapse (evident also in the t-SNE plots), where synthetic data is generated from a limited space. Similarly, for the sepsis dataset (Figure 2), CA-GAN covers more of the real data distribution compared to SMOTE, while WGAN-GP* again tends towards mode collapse.

**Fig. 1:**
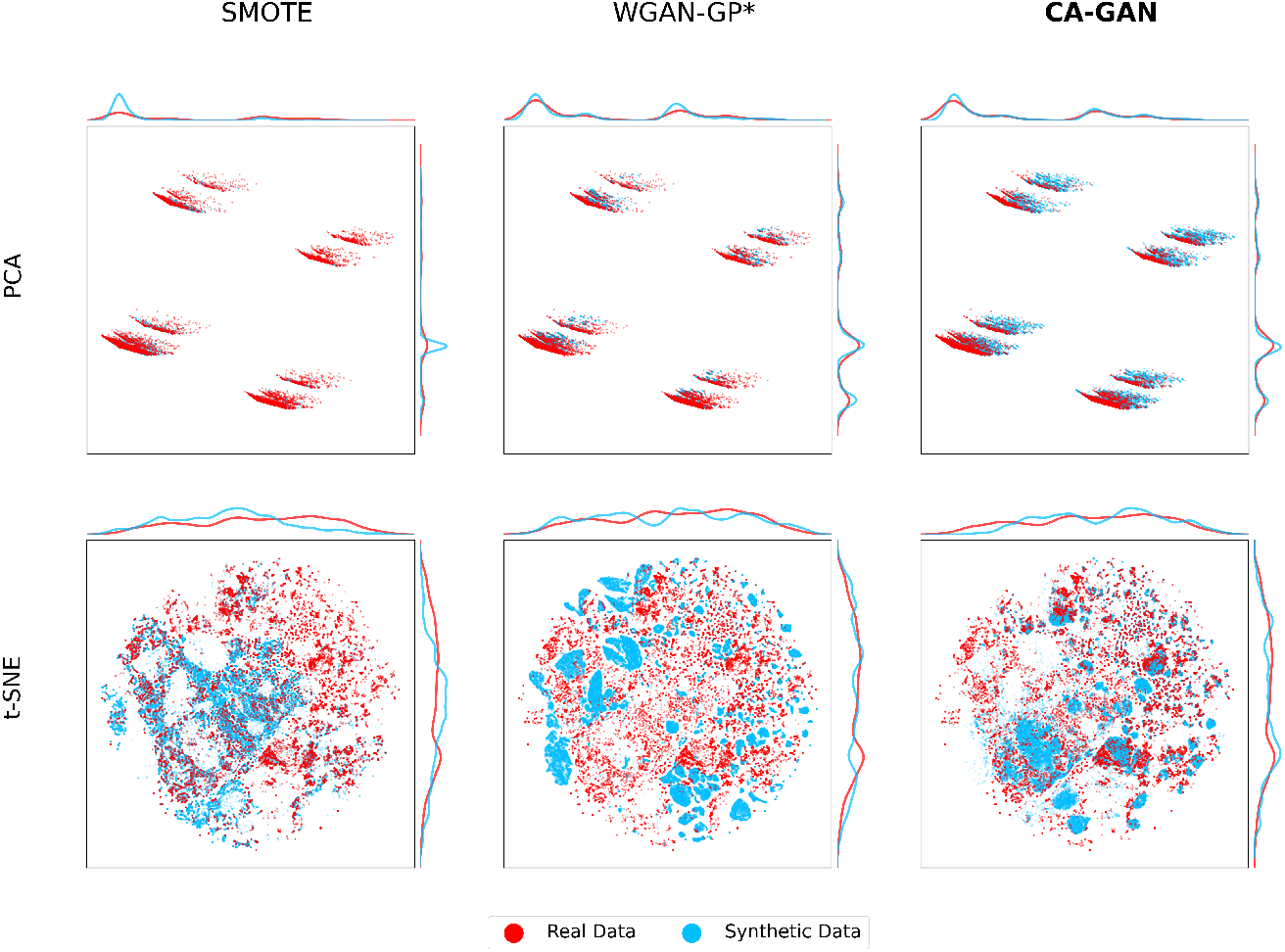
Two-dimensional representations of the acute hypotension dataset for Black patients, including marginal distributions of the principal components. Top panels: PCA two-dimensional representation of real (red) and synthetic (blue) data, where CA-GAN provides the best overall coverage of real data distribution, while SMOTE and WGAN-GP* show evidence of reduced coverage and mode collapse. Bottom panels: t-SNE two-dimensional representation of real data (red) and synthetic data (blue) for the three methods SMOTE, WGAN-GP*, and CA-GAN. It can be seen that CA-GAN more uniformly covers the real distribution, while SMOTE does not cover a significant part of it (top right in the panel) and WGAN-GP* coverage is almost completely separated from the real data.

**Fig. 2:**
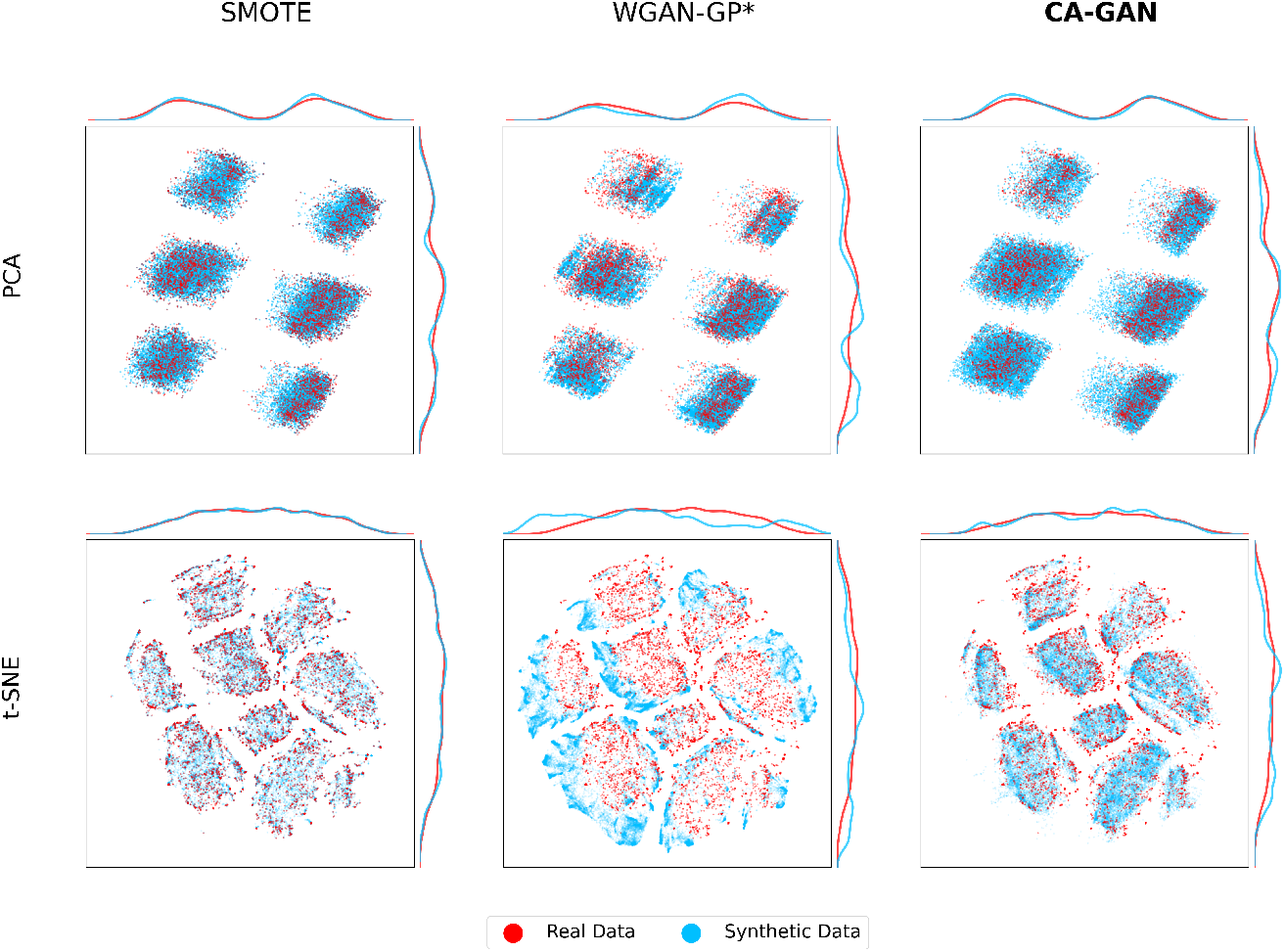
Two-dimensional representations of the sepsis dataset for Black patients, including marginal distributions of the principal components. Top panels: PCA two-dimensional representation of real (red) and synthetic (blue) data, where CA-GAN provides more coverage than SMOTE (especially in the top right and bottom left part of the panel), while WGAN-GP* provides the lowest coverage. Bottom panels: t-SNE two-dimensional representation of real data (red) and synthetic data (blue) for the three methods SMOTE, WGAN-GP*, and CA-GAN. It can be seen that SMOTE follows an interpolation pattern, while CA-GAN expands into latent space, generating authentic data points while remaining within the clusters identified by t-SNE. Data generated by WGAN-GP* fall outside of the real data.

Figures 1 and Figure 2 in the bottom panels show the t-SNE representations of the real and synthetic data. For the acute hypotension dataset, CA-GAN more uniformly covers the distribution of real data, while SMOTE does not cover a significant part of it (top right in the panel). This is also evident from the marginal distributions. WGAN-GP* coverage is almost completely separated from the real data. For the sepsis dataset, t-SNE shows that SMOTE follows an interpolation pattern failing to expand into the latent space. In contrast, CA-GAN successfully expands the distribution into the latent space, generating authentic data points, while remaining within the clusters identified by t-SNE. Data generated by WGAN-GP* fall outside of the real data.

Figures 1 and 2 provide evidence that state of the art WGAN-GP* appears to suffer from mode collapse, a significant limitation of GANs [33]. Mode collapse occurs when the generator produces a limited variety of samples despite being trained on a diverse dataset. The generator cannot fully capture the complexity of the target distribution, limiting the quantity of generated samples and resulting in repetitive output. This is because the generator can get stuck in a local minimum where a few outputs are repeatedly generated, even though the training data contains more modes that can be learned. This presents a significant challenge in generating high-quality, authentic samples, while our CA-GAN model overcomes this limitation.

The evidence that CA-GAN captures accurately the underlying structure of real data is further reinforced based on joint distribution of variables, which we show in Appendix D. In Appendix G we also present the UMAP latent representation of the data, which preserves the global structure.

Finally, we show the distribution of individual variables of synthetic data overlaid on the distribution of the real data. We use this to compare the performance of our method with state of the art WGAN-GP* as well as SMOTE as the baseline method, using acute hypotension dataset in Figure 3 and sepsis in Appendix A. Joint distributions are shown inAppendix D). The distribution of synthetic data generated by our CA-GAN exhibits the closest match to that of the real data. This close alignment is particularly evident in variables related to blood pressure, including MAP, diastolic, and systolic measurements. However, certain variables, such as urine and ALT/AST, pose challenges for all three methods. These variables have highly skewed, non-Gaussian distributions with long tails, making them difficult to transform effectively using power or logarithmic transformations. In contrast, our CA-GAN and WP-GAN* effectively capture the distribution of categorical variables. Conversely, SMOTE encounters difficulties with several variables, including both the numeric variable of urine and the categorical variable of the Glasgow Coma Score (GCS). These observations are also reflected in the quantitative evaluation in Section 2.2. The variables in the sepsis dataset are not only more than twice as many as those in acute hypotension but also have more complex distributions. Variables such as SGOT, SGPT, total bilirubin, maximum dose of vasopressors, and others have extremely long tails. The three methods struggle to generate these kinds of distributions and show a tendency to converge to the median value. In contrast, the behaviour is similar to acute hypotension for categorical and numerical variables normally distributed.

**Fig. 3:**
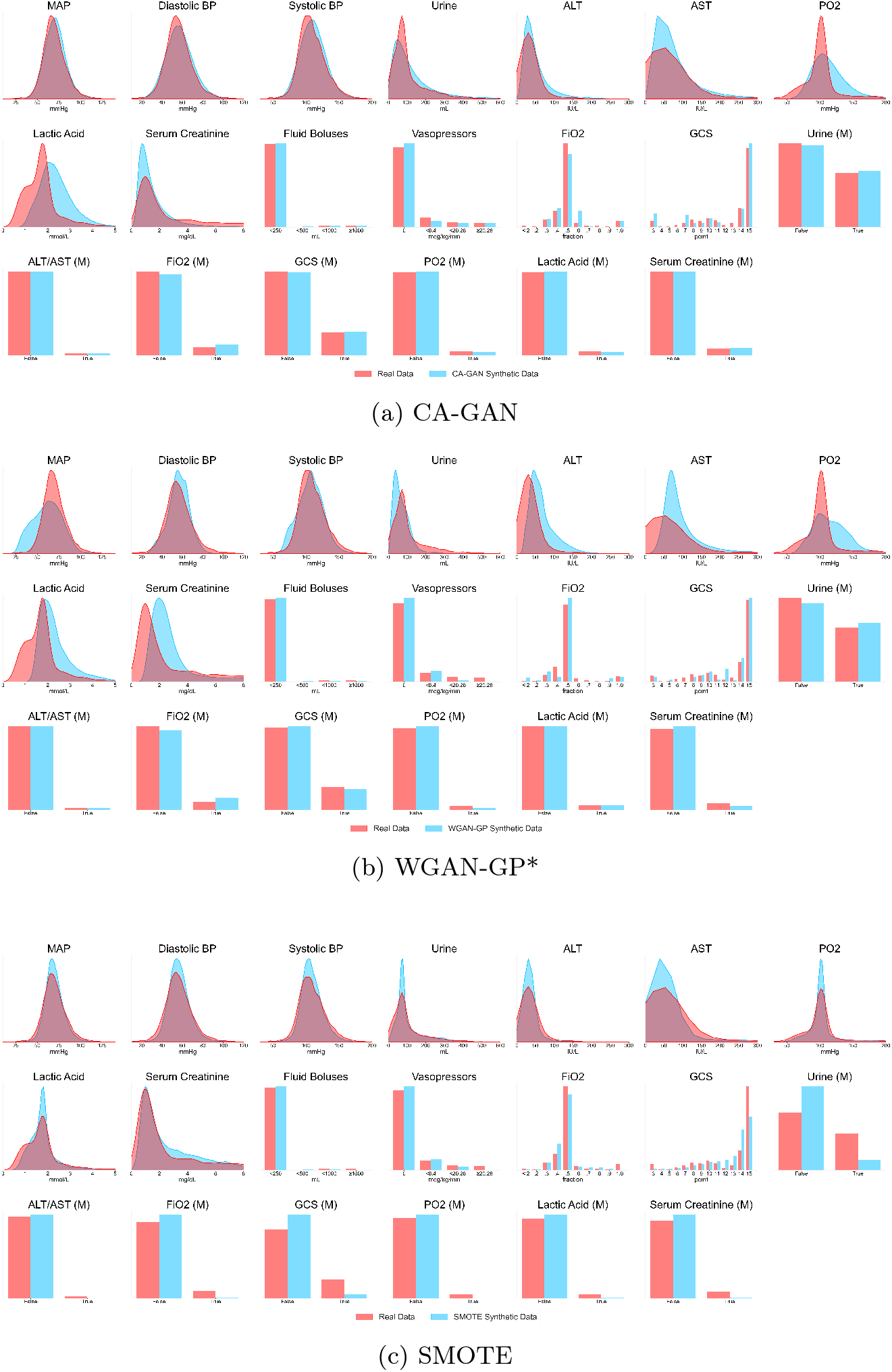
Distribution plots of each variable, overlaying real and synthetic data for acute hypotension dataset. Distribution of variables related to blood pressure (MAP, diastolic and systolic) is captured well by our method in comparison to WGAN-GP* and SMOTE. CA-GAN performs better also for categorical variables, while all the three methods struggle with variables with long tail, non-normal distributions.

### 2.2 Quantitative evaluation

We used Kullback-Leibler (KL) divergence [34] to measure the similarity between the discrete density function of the real data and that of the synthetic data. For each variable *v* of the dataset, we calculate:

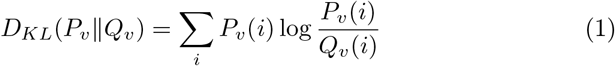

where *Q*_*v*_ is the true distribution of the variable and *P*_*v*_ is the generated distribution. The smaller the divergence, the more similar the distributions; zero indicates identical distributions. The left half of Tables 1a and 1b show the results of the KL divergence for each variable. Our CA-GAN method has the lowest median across all variables for acute hypotension and sepsis data compared to WGAN-GP* and SMOTE. This is despite the fact that SMOTE is specifically designed to maintain the distribution of the original variables.

**Table 1:** KL-Divergence and Maximum Mean Discrepancy between the distribution of real and synthetic data for each variable of the datasets.

|  | KL-divergence |  |  | MMD |  |  |
| --- | --- | --- | --- | --- | --- | --- |
|  | SMOTE | WGAN-GP* | CA-GAN | SMOTE | WGAN-GP* | CA-GAN |
| MAP | 0.11182 | 0.24941 | 0.17164 | 0.00137 | 0.00824 | 0.00110 |
| Diastolic BP | 0.28191 | 0.91622 | 0.24342 | 0.00155 | 0.00209 | 0.00086 |
| Systolic BP | 0.06405 | 0.10588 | 0.13194 | 0.00138 | 0.00120 | 0.00092 |
| Fluid Boluses | 0.01121 | 0.00358 | 0.00052 | 0.00047 | 0.00022 | 0.00003 |
| Urine | 0.00892 | 0.15183 | 0.00901 | 0.01321 | 0.08567 | 0.08443 |
| Vasopressors | 0.03622 | 0.05955 | 0.00175 | 0.00463 | 0.00883 | 0.00031 |
| ALT | 0.00068 | 0.37020 | 0.00800 | 0.01356 | 0.20156 | 0.18616 |
| AST | 0.00083 | 0.18162 | 0.00455 | 0.01323 | 0.20920 | 0.19538 |
| FiO2 | 0.00858 | 0.01950 | 1.36841 | 0.00091 | 0.00043 | 0.00012 |
| GCS | 0.02432 | 0.02571 | 0.01934 | 0.05206 | 0.00688 | 0.00791 |
| PO2 | 0.00315 | 0.13503 | 0.31726 | 0.00992 | 0.25091 | 0.24806 |
| Lactic Acid | 0.03192 | 0.42781 | 0.45402 | 0.01084 | 0.16273 | 0.19777 |
| Serum Creatinine | 0.02079 | 0.02851 | 0.08827 | 0.01892 | 0.22812 | 0.03313 |
| Urine (M) | 0.19717 | 0.00279 | 0.00070 | 0.09954 | 0.00170 | 0.00043 |
| ALT/AST (M) | 0.01872 | 0.00027 | 0.00031 | 0.00050 | 0.00001 | 0.00002 |
| FiO2 (M) | 0.07361 | 0.00965 | 0.00459 | 0.00892 | 0.00224 | 0.00103 |
| GCS (M) | 0.12043 | 0.00072 | 0.00013 | 0.03776 | 0.00030 | 0.00006 |
| PO2 (M) | 0.03846 | 0.00751 | 0.00033 | 0.00238 | 0.00067 | 0.00003 |
| Lactic Acid (M) | 0.03962 | 0.00010 | 0.00136 | 0.00274 | 0.00001 | 0.00015 |
| Serum Creatinine (M) | 0.05844 | 0.00777 | 0.00005 | 0.00613 | 0.00117 | 0.00001 |
| Median | 0.03407 | 0.02711 | **0.00629** | 0.00752 | 0.00217 | **0.00089** |

(a) Acute hypotension data
|  | KL-divergence |  |  | MMD |  |  |
| --- | --- | --- | --- | --- | --- | --- |
|  | SMOTE | WGAN-GP* | CA-GAN | SMOTE | WGAN-GP* | CA-GAN |
| Age | 0.01796 | 0.03107 | 0.02815 | 0.00543 | 0.00960 | 0.00849 |
| Heart Rate | 0.01140 | 0.07822 | 0.03885 | 0.00158 | 0.00438 | 0.00243 |
| Systolic BP | 0.12038 | 0.09645 | 0.08592 | 0.00144 | 0.00319 | 0.00262 |
| Mean BP | 0.08035 | 0.29433 | 0.27170 | 0.00192 | 0.01012 | 0.00286 |
| Diastolic BP | 0.16654 | 0.30986 | 0.39555 | 0.00186 | 0.00319 | 0.00186 |
| Respiratory Rate | 0.03584 | 0.04324 | 0.04050 | 0.00395 | 0.00273 | 0.02020 |
| Potassium | 0.15016 | 0.11200 | 0.27340 | 0.00546 | 0.01811 | 0.00742 |
| Sodium | 0.22161 | 0.18242 | 0.19199 | 0.00372 | 0.01702 | 0.00124 |
| Chloride | 0.03949 | 0.06267 | 0.00989 | 0.00291 | 0.00410 | 0.00403 |
| Calcium | 0.33564 | 0.16861 | 0.13301 | 0.00604 | 0.06938 | 0.00585 |
| Ionised Ca | 1.88754 | 0.07055 | 0.01103 | 0.00005 | 0.90096 | 0.77536 |
| CO2 | 0.04157 | 0.09045 | 0.05244 | 0.00371 | 0.00170 | 0.00118 |
| Albumin | 0.01604 | 0.00967 | 0.00532 | 0.00719 | 0.01420 | 0.00534 |
| Hemoglobin | 0.44026 | 0.30347 | 0.20345 | 0.00612 | 0.04899 | 0.02199 |
| pH | 0.12049 | 0.30020 | 0.08537 | 0.00002 | 0.00215 | 0.00003 |
| Arterial Base Excess | 0.14158 | 0.12252 | 0.04117 | 0.00410 | 0.01532 | 0.01113 |
| HCO3 | 0.02426 | 0.12058 | 0.01444 | 0.00387 | 0.01526 | 0.00256 |
| FiO2 | 0.01122 | 0.03122 | 0.03055 | 0.00022 | 0.00480 | 0.00160 |
| Glucose | 0.02167 | 0.12281 | 0.11088 | 0.00075 | 0.00041 | 0.00085 |
| Blood Urea Nitrogen | 0.04561 | 0.06180 | 0.05283 | 0.00211 | 0.00440 | 0.00391 |
| Creatinine | 0.00403 | 0.02395 | 0.13305 | 0.00739 | 0.04867 | 0.01437 |
| Magnesium | 0.05638 | 0.33564 | 0.18024 | 0.00158 | 0.04239 | 0.00214 |
| SGOT | 0.00104 | 0.00859 | 0.00199 | 0.00229 | 0.00907 | 0.00223 |
| SGPT | 0.00152 | 0.01028 | 0.00277 | 0.00198 | 0.00459 | 0.00340 |
| Total Bilirubin | 0.00493 | 0.02875 | 0.02077 | 0.00900 | 0.11124 | 0.00337 |
| WBC | 0.13135 | 0.08064 | 0.00788 | 0.00328 | 0.00573 | 0.02103 |
| Platelets Count | 0.00717 | 0.07344 | 0.00397 | 0.00116 | 0.00120 | 0.00079 |
| paO2 | 0.01067 | 0.27433 | 0.02362 | 0.00091 | 0.00321 | 0.00071 |
| paCO2 | 0.01632 | 0.05257 | 0.15250 | 0.00145 | 0.00729 | 0.00113 |
| Lactate | 0.00228 | 0.19152 | 0.11907 | 0.00829 | 0.01251 | 0.00342 |
| Input Fluids Total | 0.01047 | 0.00990 | 0.01833 | 0.00116 | 0.00208 | 0.00190 |
| Input 4H | 0.00397 | 0.00481 | 0.00471 | 0.02146 | 0.04044 | 0.02819 |
| Max Vasopressors 4H | 0.00886 | 0.00240 | 0.00295 | 0.00028 | 0.00083 | 0.00133 |
| Total Urine Output | 0.01397 | 0.01009 | 0.00832 | 0.00395 | 0.01644 | 0.01511 |
| Output 4H | 0.03328 | 0.00800 | 0.00484 | 0.00947 | 0.04256 | 0.02401 |
| Gender | 0.00117 | 0.02196 | 0.01476 | 0.01363 | 0.01384 | 0.00931 |
| Readmission of Patient | 0.00091 | 0.02287 | 0.01917 | 0.00064 | 0.01346 | 0.01134 |
| Mechanical Ventilation | 0.00065 | 0.02689 | 0.00577 | 0.00054 | 0.01685 | 0.00364 |
| Temperature | 0.00035 | 0.01592 | 0.00722 | 0.00011 | 0.00336 | 0.00236 |
| GCS | 0.01751 | 0.06029 | 0.01191 | 0.02948 | 0.02881 | 0.00903 |
| SpO2 | 0.03123 | 0.09465 | 0.06963 | 0.02013 | 0.04669 | 0.03336 |
| PTT | 0.02686 | 0.03667 | 0.02221 | 0.01405 | 0.01906 | 0.01723 |
| PT | 0.03676 | 0.03933 | 0.00986 | 0.02061 | 0.03367 | 0.00594 |
| INR | 0.02486 | 0.03850 | 0.00669 | 0.02113 | 0.01912 | 0.00568 |
| Median | 0.02296 | 0.06105 | **0.02292** | **0.00349** | 0.01299 | 0.00377 |

(b) Sepsis data

In addition, we used Maximum Mean Discrepancy (MMD) [35] to calculate the distance between the distributions based on kernel embeddings, that is, the distance of the distributions represented as elements of a reproducing kernel Hilbert space (RKHS). We used a Radial Basis Function (RBF) Kernel:

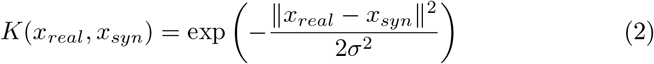

with *σ* = 1. The right half of Tables 1a and 1b shows the MMD results for SMOTE, WGAN-GP* and our CA-GAN. Again, our model has the best median performance across all the variables for acute hypotension data, while for sepsis data, SMOTE shows a difference in performance by 0.00028. In summary, CA-GAN performs best in the acute hypotension dataset by a wide margin while showing comparable performance with SMOTE in the sepsis dataset. A summary of distance metrics, including median, mean, standard deviation, maximum and minimum is shown in Appendix E.

### 2.3 Variable correlations

We used the Kendall rank correlation coefficient *τ* [36] to investigate whether synthetic data maintained original correlations between variables found in the real data of acute hypotension and sepsis datasets. This choice is motivated by the fact that the *τ* coefficient does not assume a normal distribution, which is the case for some of our variables, of the sepsis dataset in particular (as shown in Figure 3 and Appendix A). Figure 4 shows the results of Kendall’s rank correlation coefficients. For the acute hypotension dataset (Figure 4a), CA-GAN captures the original variable correlations, as does SMOTE, with the former having the closest results on categorical variables, while the latter on numerical ones. WGAN-GP* shows the worst performance, accentuating correlations that do not exist in real data. Similar patterns are also obtained for the variables of patients with sepsis in Figure 4b. For additional insight, Appendix H presents the absolute difference between correlations, highlighting the same patterns.

**Fig. 4:**
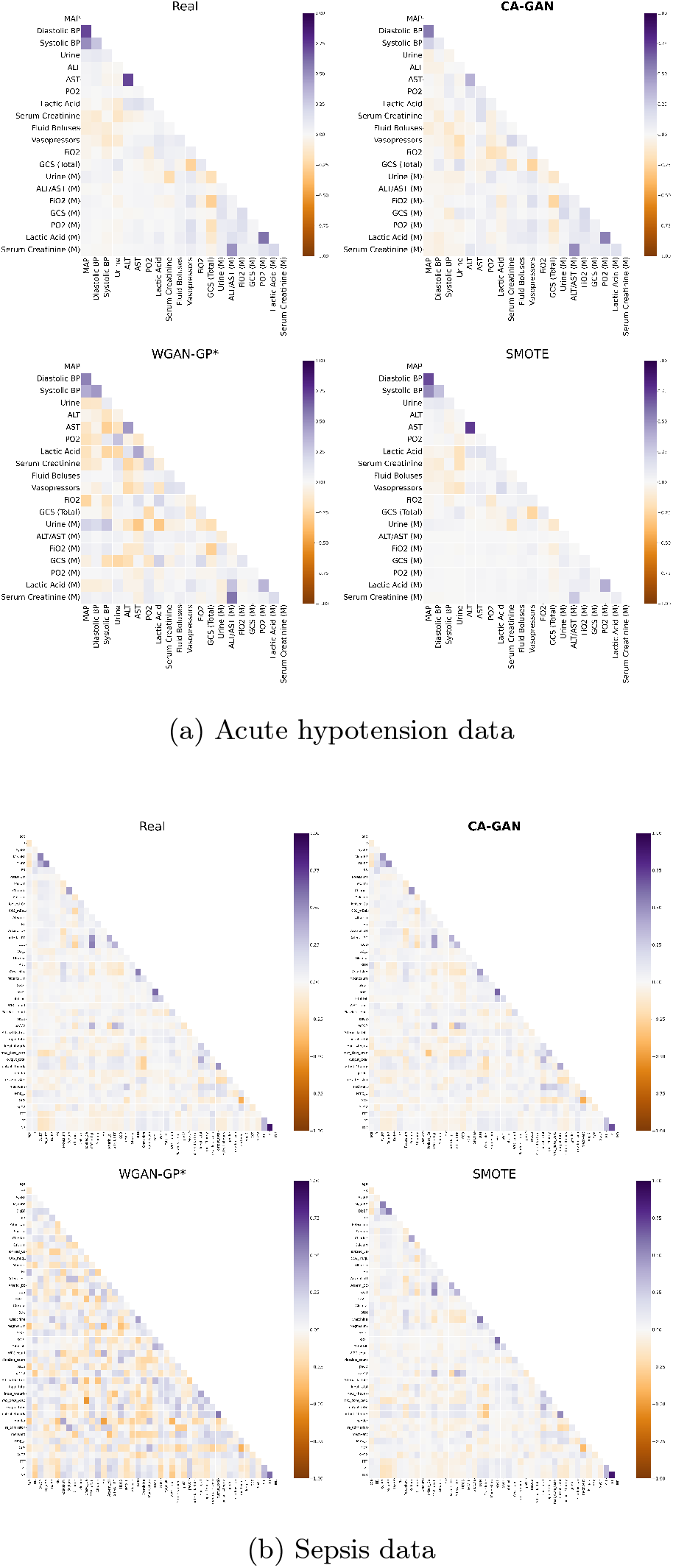
Kendall’s rank correlation coefficients for the real data and the data generated with CA-GAN, WGAN-GP*, and SMOTE.

### 2.4 Synthetic data authenticity

When generating synthetic data, the output must be a realistic representation of the original data. Still, we also need to verify that the model has not merely learned to copy the real data. GANs are prone to overfitting by memorising the real data [37]; therefore, we use Euclidean Distance (*L*_2_ Norm) to evaluate the originality of our model’s output. Our analysis shows that the smallest distance between a synthetic and a real sample is 52.6 for acute hypotension and 44.2 for sepsis, indicating that the generated synthetic data are not a mere copy of the real data. This result, coupled with the visual representation of CA-GAN (shown in Figure 1 and 2), illustrates the ability of our model to generate authentic data. SMOTE, which by design interpolates the original data points, is unable to explore the underlying multidimensional space. Therefore its generated data samples are much closer to the real ones, with a minimum Euclidean distance of 0.0023 for acute hypotension and 0.033 for sepsis. Table 2 shows a summary of the minimum Euclidian distance for each method.

**Table 2:** Minimum Euclidean distance between real and synthetic data generated by SMOTE, WGAN-GP* and CA-GAN. No method generates exact copies of the real data.

|  | SMOTE | WGAN-GP* | CA-GAN |
| --- | --- | --- | --- |
| Acute Hypotension | 0.0023 | 65.95 | 52.56 |
| Sepsis | 0.033 | 47.91 | 44.18 |

### 2.5 Improving model fairness

Having evaluated the quality of synthetic data generated by our CA-GAN, we now focus on evaluating the potential of synthetic data in improving model fairness. The machine learning literature has highlighted many examples in which class imbalanced datasets lead to unfair decisions for the minority class [38]. This is especially important when those decisions significantly affect the health of patients and the society as a whole, as is the case with our study. We measure fairness by comparing the performance of the model between subgroups, based on the approach described in [39]. In this respect, we have performed an analysis to understand the impact of synthetic data in the performance of a predictive model within Black patients and White patients separately. For this purpose, we chose the task of predicting blood lactate within each ethnic subgroup, based on our previous work [40]. Blood lactate is essential in guiding clinical decision-making, especially for patients with sepsis and ultimately affects patients’ survival [41, 42]. Therefore, any differences between Black and White patients in lactate prediction performance would result in potentially unfair treatment decisions due to data representation bias.

Based on our work in [40] we trained a Random Forest classifier to predict whether the outcome of the last lactate value in the time series of patients was above a clinically relevant threshold, using as input the previous observations of the clinical variables in our dataset. Initially, we devised a fixed test set of real data, that was not seen by our generative model and we ran our generative model five times. We used only the real (original) sepsis dataset where the predictive performance of the classifier within the Black patient cohort was AUC of 0.569 (*±* 0.020). This is in contrast to AUC of 0.652 (*±* 0.016) within the White patient cohort. For comparison, the overall performance of the model when including both ethnicities was AUC of 0.611 (*±* 0.013). Then we augmented the original Black patient cohort with synthetic data (conditioned on Black patients only) generated by our CA-GAN model, such that the representation between Black and White patients was equal. As a consequence, the predictive performance within Black patient cohort increased to AUC of 0.620 (*±* 0.017), while for White patients it was AUC of 0.643 (*±* 0.014) and AUC of 0.629 (*±* 0.013) for the overall test set with both ethnicities.

Overall, synthetic data augmentation resulted in a fairer model between ethnicities. The performance gap between ethnicities was reduced from 8% (ΔAUC = 0.083 *±*0.028) to 2% (ΔAUC = 0.025 *±* 0.015), with a statistically significant difference between non-augmented and augmented datasets (p=0.0236).

### 2.6 Downstream regression task on ethnicity

Finally, we also sought to evaluate the ability of CA-GAN to maintain the temporal properties of time series data, considering ethnicity and gender (in the following section). Since our objective is to augment the minority class to mitigate representation bias, we wanted to verify that the datasets augmented with synthetic data generated by our model can maintain or improve the predictive performance of the original data on a downstream task. Initially, we trained only a Bidirectional Long Short-Term Memory (BiLSTM) with real data as the baseline. Later, we trained the BiLSTM with synthetic and augmented datasets separately, considering ethnicity and gender diversity. They were used to evaluate the performance in a regression task. To address the inherent randomness in the models, we used CA-GAN to create five different synthetic datasets. For each of these datasets, we trained five BiLSTM models, resulting in a total of 25 BiLSTM models (5 datasets × 5 models per dataset). A complete description of how we performed these experiments is provided in Appendix F.

Table 3a and Table 3b show the mean absolute errors between the BiLSTM prediction and the actual acute hypotension and sepsis observations, respectively. In these experiments, the test sets consisted exclusively of patients from the minority class. In the first column, we show the results achieved using only the real data to make the predictions; in the second column, the results using only the synthetic data; and finally, in the third column, the results achieved by predicting with the augmented dataset, that is, with both the real and synthetic data together, considering both ethnic and gender diversity in the augmentation process. Overall, adding the synthetic data reduces the predictive error. This indicates that the temporal characteristics of the data generated by our CA-GAN model are close enough to those of the real data to maintain the original predictive performance. Thus, the augmented dataset could be used in a downstream task, mitigating the representation bias.

**Table 3:** Mean prediction errors of a BiLSTM trained on real, synthetic, and augmented data for a downstream prediction task. The numbers in parentheses represent the standard deviation

|  | Real | Synthetic | Augmented |
| --- | --- | --- | --- |
| MAP | 10.129 (0.487) | 9.084 (0.401) | 8.530 (0.234) |
| Diastolic BP | 8.328 (0.653) | 7.500 (0.246) | 7.491 (0.205) |
| Systolic BP | 19.930 (0.752) | 16.375 (1.405) | 13.121 (0.646) |
| Fluid Boluses | 19.547 (2.387) | 22.987 (2.051) | 22.684 (1.544) |
| Urine | 54.915 (1.591) | 49.615 (2.201) | 49.461 (3.279) |
| Vasopressors | 0.290 (0.051) | 0.446 (0.079) | 0.430 (0.082) |
| ALT | 7.561 (0.616) | 8.244 (1.079) | 6.235 (2.018) |
| AST | 18.017 (3.876) | 9.629 (2.106) | 8.091 (2.377) |
| FIO2 | 0.022 (0.019) | 0.023 (0.008) | 0.028 (0.015) |
| GCS | 0.226 (0.069) | 0.481 (0.093) | 0.666 (0.177) |
| PO2 | 26.745 (9.310) | 12.297 (1.184) | 11.375 (4.392) |
| Lactic Acid | 0.107 (0.064) | 0.262 (0.018) | 0.168 (0.023) |
| Serum Creatinine | 0.158 (0.054) | 0.357 (0.037) | 0.321 (0.040) |
| Median | 8.493 (0.639) | 8.255 (0.731) | **6.844** (0.843) |

(a) Acute hypotension data
|  | Real | Synthetic | Augmented |
| --- | --- | --- | --- |
| Heart Rate | 13.926 (1.406) | 6.462 (0.510) | 6.061 (0.238) |
| Systolic BP | 20.975 (0.411) | 12.129 (1.048) | 11.636 (0.704) |
| Mean BP | 12.628 (0.885) | 7.706 (0.376) | 7.447 (0.458) |
| Diastolic BP | 9.793 (0.468) | 8.427 (0.286) | 8.282 (0.446) |
| Respiratory Rate | 2.515 (0.270) | 2.388 (0.065) | 2.341 (0.039) |
| Potassium | 0.183 (0.020) | 0.207 (0.027) | 0.220 (0.048) |
| Sodium | 28.981 (1.978) | 3.698 (0.077) | 3.733 (0.122) |
| Chloride | 8.125 (0.396) | 4.533 (0.368) | 4.080 (0.483) |
| Calcium | 0.388 (0.043) | 0.327 (0.057) | 0.388 (0.118) |
| Ionised Ca | 0.052 (0.001) | 0.092 (0.043) | 0.086 (0.041) |
| Carbon Dioxide | 2.324 (0.206) | 2.033 (0.177) | 1.893 (0.158) |
| Albumin | 0.516 (0.006) | 0.452 (0.020) | 0.442 (0.008) |
| Hemoglobin | 0.725 (0.087) | 0.616 (0.063) | 0.655 (0.079) |
| pH | 0.082 (0.050) | 0.083 (0.022) | 0.075 (0.041) |
| Arterial Base Excess | 2.972 (0.070) | 2.876 (0.019) | 2.847 (0.019) |
| HCO3 | 3.244 (0.628) | 2.391 (0.065) | 2.452 (0.142) |
| FIO2 | 0.068 (0.025) | 0.063 (0.016) | 0.046 (0.014) |
| Glucose | 34.395 (0.765) | 24.366 (0.543) | 23.388 (0.765) |
| Blood Urea Nitrogen | 4.096 (0.765) | 4.484 (0.429) | 3.430 (0.343) |
| Creatinine | 0.135 (0.035) | 0.257 (0.056) | 0.276 (0.041) |
| Magnesium | 0.109 (0.015) | 0.126 (0.023) | 0.126 (0.031) |
| SGOT | 216.348 (2.019) | 206.530 (0.731) | 207.164 (1.537) |
| SGPT | 165.979 (1.735) | 153.379 (1.475) | 153.672 (1.527) |
| Total Bilirubin | 1.550 (0.089) | 1.457 (0.155) | 1.398 (0.028) |
| WBC | 1.072 (0.208) | 1.516 (0.144) | 1.264 (0.097) |
| Platelets Count | 119.777 (4.692) | 59.953 (4.340) | 62.003 (6.094) |
| paO2 | 37.749 (0.064) | 35.157 (1.201) | 35.318 (1.989) |
| paCO2 | 7.206 (0.454) | 6.156 (0.377) | 6.035 (0.292) |
| Lactate | 1.026 (0.174) | 0.864 (0.024) | 0.860 (0.018) |
| Input Fluids Total | 9824.288 (6.460) | 9378.006 (56.594) | 9354.273 (27.237) |
| Input 4H | 195.774 (1.827) | 177.704 (1.970) | 178.058 (4.157) |
| Max Vasopressors 4H | 0.048 (0.018) | 0.085 (0.043) | 0.126 (0.144) |
| Total Urine Output | 9067.905 (4.940) | 8738.295 (16.576) | 8770.296 (29.386) |
| Output 4H | 235.173 (4.450) | 169.318 (5.368) | 168.216 (4.849) |
| Median | 3.932 (0.536) | 2.876 (0.019) | **2.847** (0.019) |

(b) Sepsis data

**Table 4:** Results of downstream regression task on gender-conditioned data, based on Mean Absolute Error (MAE), with standard deviation shown in brackets. Biased Real represents the real datatset from which we have removed 80% of the data from female patients.

|  | Biased Real | Synthetic | Real | Augmented |
| --- | --- | --- | --- | --- |
| Heart Rate | 12.095 (0.190) | 6.636 (0.536) | 6.526 (0.330) | 6.182 (0.250) |
| Systolic BP | 21.662 (1.290) | 10.803 (0.896) | 11.453 (0.549) | 9.909 (0.387) |
| Mean BP | 10.280 (0.054) | 6.342 (0.201) | 7.055 (0.587) | 6.507 (0.287) |
| Diastolic BP | 9.308 (0.666) | 6.304 (0.186) | 7.029 (0.406) | 6.131 (0.049) |
| Respiratory Rate | 2.461 (0.071) | 2.478 (0.196) | 2.444 (0.065) | 2.419 (0.107) |
| Potassium | 0.258 (0.047) | 0.195 (0.012) | 0.158 (0.019) | 0.190 (0.027) |
| Sodium | 32.669 (1.528) | 3.793 (0.149) | 3.653 (0.071) | 3.891 (0.204) |
| Chloride | 10.612 (1.388) | 4.013 (0.561) | 5.013 (0.240) | 3.700 (1.495) |
| Calcium | 0.425 (0.030) | 0.537 (0.129) | 0.190 (0.060) | 0.411 (0.091) |
| Ionised Ca | 0.070 (0.015) | 0.110 (0.032) | 0.056 (0.004) | 0.101 (0.043) |
| Carbon Dioxide | 3.034 (0.820) | 2.462 (0.155) | 2.667 (0.744) | 2.306 (0.112) |
| Albumin | 0.514 (0.045) | 0.458 (0.009) | 0.474 (0.010) | 0.460 (0.013) |
| Hemoglobin | 0.729 (0.066) | 0.677 (0.153) | 0.596 (0.021) | 0.666 (0.086) |
| pH | 0.070 (0.020) | 0.073 (0.044) | 0.076 (0.038) | 0.082 (0.019) |
| Arterial Base Excess | 2.632 (0.098) | 2.612 (0.037) | 2.605 (0.078) | 2.547 (0.032) |
| HCO3 | 3.278 (0.293) | 2.651 (0.136) | 3.172 (0.571) | 2.490 (0.048) |
| FiO2 | 0.075 (0.015) | 0.064 (0.003) | 0.058 (0.003) | 0.066 (0.005) |
| Glucose | 46.868 (0.555) | 28.239 (3.333) | 24.431 (2.191) | 26.405 (1.246) |
| Blood Urea Nitrogen | 6.425 (0.814) | 5.449 (0.414) | 3.500 (0.979) | 4.583 (0.329) |
| Creatinine | 0.126 (0.025) | 0.252 (0.076) | 0.172 (0.107) | 0.169 (0.022) |
| Magnesium | 0.111 (0.015) | 0.135 (0.041) | 0.096 (0.021) | 0.124 (0.028) |
| SGOT | 130.591 (1.089) | 127.034 (1.791) | 136.685 (3.285) | 126.824 (2.759) |
| SGPT | 112.892 (2.177) | 113.651 (5.488) | 118.709 (3.904) | 112.270 (3.293) |
| Total Bilirubin | 1.552 (0.076) | 1.347 (0.147) | 1.552 (0.252) | 1.398 (0.084) |
| WBC | 0.990 (0.221) | 1.280 (0.297) | 1.101 (0.612) | 1.195 (0.305) |
| Platelets Count | 134.399 (4.411) | 53.030 (3.271) | 43.874 (4.875) | 56.936 (5.577) |
| paO2 | 45.729 (0.515) | 38.913 (0.811) | 42.085 (1.316) | 38.054 (1.368) |
| paCO2 | 6.954 (0.195) | 5.867 (0.165) | 6.866 (1.254) | 5.861 (0.131) |
| Lactate | 0.796 (0.048) | 0.725 (0.015) | 0.759 (0.025) | 0.734 (0.020) |
| Input Fluids Total | 10524.103 (9.990) | 10136.382 (24.197) | 10325.518 (27.476) | 10086.376 (51.517) |
| Input 4H | 158.433 (1.912) | 137.386 (1.772) | 154.167 (5.623) | 138.082 (1.460) |
| Max Vasopressors 4H | 0.065 (0.033) | 0.036 (0.010) | 0.026 (0.016) | 0.033 (0.007) |
| Total Urine Output | 10124.596 (5.131) | 9774.791 (31.050) | 9981.963 (13.985) | 9759.179 (22.349) |
| Output 4H | 210.089 (11.468) | 159.417 (3.038) | 163.217 (3.371) | 158.430 (3.308) |
| Median | 4.784 (0.393) | 3.193 (0.052) | 3.294 (0.241) | **2.870** (0.220) |

It should be noted that the errors in Input Fluids Total and Total Urine Output are exceptionally high compared to the other variables. This is because predicting these variables is generally challenging, stemming partly from how they are collected and recorded rather than an issue inherent to synthetic data generation.

### 2.7 Downstream regression task on gender-conditioned data

As a final test, we have devised an additional dataset of patient subpopulations with an artificially biased gender representation. Specifically, from the original sepsis data (defined as “Real”), we have removed 80% of the data from female patients to introduce a gender-based bias (“Biased Real” dataset). We used the biased dataset to train our architecture and generate synthetic data. To evaluate the performance of our approach, we applied a downstream regression task similarly to the one presented above for the ethnicity data. A test set consisting of patients from the minority class (i.e. female patients) was kept separate for this evaluation.

Our results show that the performance of data generated by our model is comparable to that of real data. Namely using the original dataset, we obtain a Mean Absolute Error (MAE) of 4.784 (±0.393), while we obtain an MAE of 3.193 (±0.052) using the synthetic dataset, indicating that the CA-GAN generates faithful synthetic data. What is even more interesting is that combining synthetic data with the real data further reduced the MAE to 2.870 (±0.220), which is lower than that of the real data alone (MAE of 3.294 (±0.241)).

## 3 Discussion

As machine intelligence scales upwards in clinical decision-making, the risk of perpetuating existing health inequities increases significantly. This is because biased decision-making can continuously feed back the data used to train the models, creating a vicious circle that further ingrains discrimination towards underrepresented groups. Representation bias, in particular, frequently occurs in health data, leading to decisions that may not be in the best interests of all patients, favouring specific subpopulations while treating underrepresented subpopulations, such as those with standard set characteristics including ethnicity, gender, and disability unfavourably.

To address these issues, representation must be improved before algorithmic decision-making becomes integral to clinical practice. While unequal representation is a multifaceted challenge involving diverse factors such as socio-economic, cultural, systemic, and data, our work represents a step towards addressing one significant facet of this challenge: mitigating existing representation bias in health data.

We have shown that our work can generate high-quality synthetic data when evaluated against state-of-the-art architectures and traditional approaches such as SMOTE. SMOTE has notable advantages over other data generation techniques as it requires no training and can work with smaller datasets. It can mirror non-normal distributions even if it tends to overestimate the median in long-tail distributions. This is in contrast to GANs, which struggle with these types of distributions. However, generating authentic data remains a significant challenge for SMOTE, especially important when considering confidentiality of data and patients’ privacy.

Through qualitative and quantitative evaluation, we have shown that CA-GAN can generate authentic data samples with high distribution coverage, avoiding mode collapse failure, while ensuring that the generated data are not copies of the real data. We have also shown that augmenting the dataset with the synthetic data generated by CA-GAN leads to lower errors in the downstream regression task. This indicates that our model can generalise well from the original data.

A notable advantage of our approach is that it uses the overall dataset, and not only the minority class, as is the case with WGAN-GP* and SMOTE. This means that CA-GAN can be applied in smaller datasets and those with highly imbalanced classes, such as rare diseases.

Furthermore, we evaluated our method on two datasets with diverse characteristics and found that our CA-GAN performed better on the acute hypotension dataset. This may be because some of the numerical variables in the sepsis dataset have long-tailed distributions, presenting a modelling challenge for all the methods. Similar challenges are observed for variables with non-normal distributions. Additionally, the sepsis dataset contained fewer data points per patient than acute hypotension (15 versus 48 observations). A shorter input sequence may have created difficulty for BiLSTM modules to learn the underlying structure of the original data effectively, coupled with a higher number of variables (twice as many) in the sepsis dataset. We also note that the lower number of patients in the acute hypotension dataset does not impact the generative performance of our method. This ability to work with fewer data points (patients) is encouraging, given the overall objective of our goal of augmenting representation.

The task of generalising to unseen data categories is particularly challenging due to the inherent unknowns these categories represent. While a definitive strategy is yet to be developed, we believe that integrating conditional generation with metric learning, as seen in prototypical networks [43], could provide a dual advantage. This integration could not only facilitate the generation of novel data points but also offer a quantifiable framework to assess their similarity or dissimilarity to known data categories. Such an approach could extend the capabilities of CA-GAN beyond data augmentation, potentially improving the interpretability and applicability of the synthetic data generated.

A future development of this work involves the use our architecture to study counterfactual examples in downstream tasks [44]. Counterfactuals would make it possible to identify specific conditions that produce disparities in model prediction, improving the understanding of bias mitigation.

In terms of evaluation metrics, our current approach primarily focuses on statistical properties of the generated data. However, to ascertain the practical utility and accuracy of the synthetic data produced by CA-GAN, we recognise the importance of domain-specific validations, including performance in subpopulations [45, 46]. Inspired by the collaborative efforts outlined in [47], we are committed to exploring partnerships with experts in relevant fields such as healthcare and clinical practice to guide the development of evaluation metrics. Their input will ensure that our synthetic datasets can meet the rigorous demands of real-world applications and contribute meaningfully to the domains they are intended to serve.

Our architecture can provide a solid basis to generate privacy preserving synthetic data and mitigate barriers to access clinical data. This is because, we have ensured that the synthetic data generated by our CA-GAN are not a mere copy of the real data on one hand, while on the other, the synthetic data reflect the distributions of the real data. However, privacy preserving aspects will require additional analysis, such as reconstruction attacks, which are beyond the scope of the current work.

While CA-GAN architecture showed superior performance with respect to state of the art method as well as computationally inexpensive approaches, some limitations are present. Namely, CA-GAN may require additional optimisations to further increase performance on datasets with variables with non Gaussian distributions and those with long-tailed distributions. Furthermore, additional analysis will be required to evaluate the generalisation capability of our architecture with datasets of different characteristics. In this respect we aim to refine CA-GAN while exploring alternative architectures (such as Diffusion Models) in addressing some of these limitations. One approach might be using convolutional neural networks (CNNs) or Temporal Convolutional Networks (TCNs) as a promising direction to potentially improve the efficiency of our model. The work of Bai et al. [48] provides an empirical foundation for this approach, indicating that such network structures can rival the performance of recurrent networks for sequence modelling tasks. Additionally, prior research [49] suggests that simplifying the internal mechanisms of these recurrent units can lead to improvements in both performance and computational efficiency. These insights provide a strong impetus for our future work, where we aim to refine the CA-GAN model to harness the benefits of these alternative architectures without compromising its ability to perform conditional generation.

Finally, we are aware that the use of synthetic data may generate several ethical and policy implications including the fact that synthetic data cannot fully address the historical biases and discriminatory practices which are often reflected in the data [50]. While our work can mitigate existing representation biases, we must ensure that this does not come at the risk of disincentivising participation of underrepresented groups or perpetuating other types of data biases [51, 52]. Finally, while we showed the utility of synthetic data, we also note that the findings should always be confirmed using real data.

## 4 Methods

We begin by formally formulating the problem we are addressing. Then we discuss the data sources we used to train our models and compare and contrast Generative Adversarial Networks (GANs) and Conditional Generative Adversarial Networks (CGANs). We also provide an in-depth analysis of the baseline model for this work, WGAN-GP*. Finally, we present the architecture of our proposed Conditional Augmentation GAN (CA-GAN) and discuss its advantages over other methods.

### 4.1 Problem Formulation

Let *A* be a vector space of features and let *a* ∈ *A* represent a feature vector. Let *L* = *{*0, 1*}* be a binary distribution modifier, and let *l* a binary mask extracted from *L*. We consider a data set 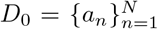 with *l* = 0, where individual samples are indexed by *n* ∈ {1, …, *N*} and we also consider a data set 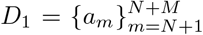 with *l* = 1, where individual samples are indexed by *m* ∈ {*N* + 1, …, *N* + *M*}, and *N > M*. We define the training data set *D* as *D* = *D*_0_ ∪ *D*_1_.

Our goal is to learn a density function 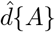 that approximates the true distribution *d* {*A*} of *D*. We also define 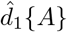 as 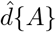 with *l* = 1 applied.

To balance the number of samples in *D*, we draw random variables *X* from 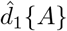 and add them to *D*_1_ until *N* = *M*. Thus, we balance out *D*.

### 4.2 Data sources, variables and patient population

Our analysis uses two datasets extracted from the MIMIC-III database. The detailed data pre-processing steps are outlined in our previous publication [27], in Section 1.2 and Section 1.3 of the supplementary material. We chose these two datasets as they have already been used in the study describing WGAN-GP* [27] making the comparison with our method fairer. We decided to test the methods for the oversampling of only one minority class, thus including only patients that belonged to the White (coded as Caucasian) or Black ethnic groups. We used a similar approach for gender, shown in Subsection 2.7.

The acute hypotension dataset comprises 3343 patients admitted to critical care; the patients were either of Black (395) or White (2948) ethnicity. Each patient is represented by 48 data points, corresponding to the first 48 hours after the admission, and 20 variables, namely nine numeric, four categorical, and seven binary variables. Details of this dataset are presented in Appendix B. The Sepsis dataset comprises 4192 patients admitted to critical care of either Black (461) or White (3731) ethnicity. Each patient is represented by 15 data points, corresponding to observations taken every four hours from admission, and 44 variables, namely 35 numeric, six categorical, and three binary variables. Details of this dataset are presented in Appendix B.

### 4.3 GAN vs CGAN

The Generative Adversarial Network (GAN) [53] entails two components: a generator and a discriminator. The generator *G* is fed a noise vector *z* taken from a latent distribution *p*_*z*_ and outputs a sample of synthetic data. The discriminator *D* inputs either fake samples created by the generator or real samples *x* taken from the true data distribution *p*_*data*_. Hence, the GAN can be represented by the following minimax loss function:

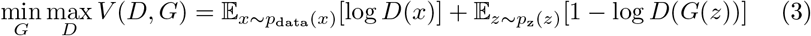

The goal of the discriminator is to maximise the probability of discerning fake from real data, whilst the purpose of the generator is to make samples realistic enough to fool the discriminator, i.e., to minimise 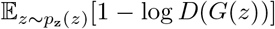. As a result of the reciprocal competition, both the generator and discriminator improve during training.

The limitations of vanilla GAN models become evident when working with highly imbalanced datasets, where there might not be sufficient samples to train the models to generate minority-class samples. A modified version of GAN, the Conditional GAN [54], solves this problem using labels *y* in both the generator and discriminator. The additional information *y* divides the generation and the discrimination in different classes. Hence, the model can now be trained on the whole dataset to generate only minority-class samples. Thus, the loss function is modified as follows:

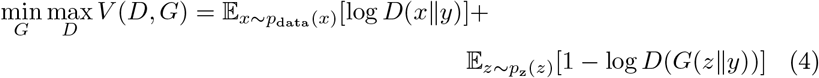

GAN and CGAN, overall, share the same significant weaknesses during training, namely mode collapse and vanishing gradient [33]. In addition, as GANs were initially designed to generate images, they have been shown unsuitable for generating time-series [55] and discrete data samples [56].

### 4.4 WGAN-GP*

The WGAN-GP* introduced by Kuo et al. [27] solved many of the limitations of vanilla GANs. The model was a modified version of a WGAN-GP [25, 26]; thus, it applied the Earth Mover distance (EM) [57] to the distributions, which had been shown to solve both vanishing gradient and mode collapse [58]. In addition, the model applied the Gradient Penalty during training, which helped to enforce the Lipschitz constraint on the discriminator efficiently. In contrast with vanilla WGAN-GP, WGAN-GP* employed soft embeddings [59, 60], which allowed the model to use inputs as numeric vectors for both binary and categorical variables, and a Bidirectional LSTM layer [61, 62], which allowed for the generation of samples in time-series. While *L*_*D*_ was kept the same, *L*_*G*_ was modified by Kuo et al. [27] by introducing alignment loss, which helped the model to capture correlation among variables over time better. Hence, the loss functions of WGAN-GP* are the following:

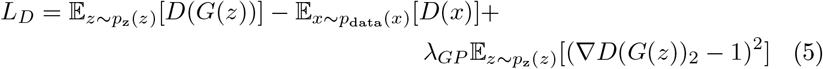

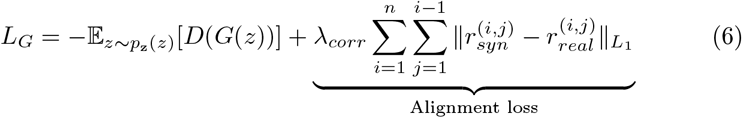

To calculate alignment loss, we computed Pearson’s r correlation [63] for every unique pair of variables *X*^(*i*)^ and *X*^(*j*)^. We then applied the *L*_1_ loss to the differences in the correlations between *r*_*syn*_ and *r*_*real*_, with *λ*_*corr*_ representing a constant acting as a strength regulator of the loss.

In their follow-up papers, Kuo et al. noted that their simulated data based on their proposed WGAN-GP* lacked diversity. In [64], the authors found that WGAN-GP* continued to suffer from mode collapse like the vanilla GAN. Similar to our own CA-GAN, the authors extended the WGAN-GP setup with a conditional element where they externally stored features of the real data during training and replayed them to the generator sub-network at test time. In [65], the same panel of researchers also experimented with diffusion models [66] and found that diffusion models better represent binary and categorical variables. Nonetheless, they demonstrated that GAN-based models encoded less bias (in the means and variances) of the numeric variable distributions.

### 4.5 CA-GAN

We built our CA-GAN by conditioning the generator and the discriminator on static labels *y*. Hence, the updated loss functions used by our model are as follows:

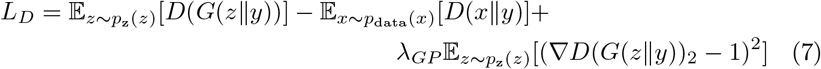

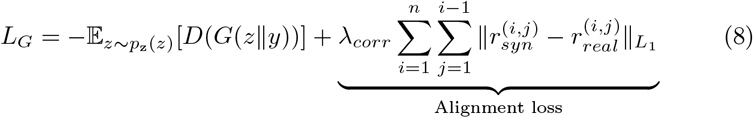

Where *y* can be any categorical label. During training, the label *y* was used to differentiate the minority from the majority class, and during generation, they were used to create fake samples of the minority class.

Compared to WGAN-GP* we also increased the number of BiLSTMs from 1 to 3 both in the generator and the discriminator, as stacked BiLSTMs have been shown to capture complex time-series better [67]. In addition, we decreased the learning rate and batch size during training. An overview of the CA-GAN architecture is shown in Figure 5.

**Fig. 5:**
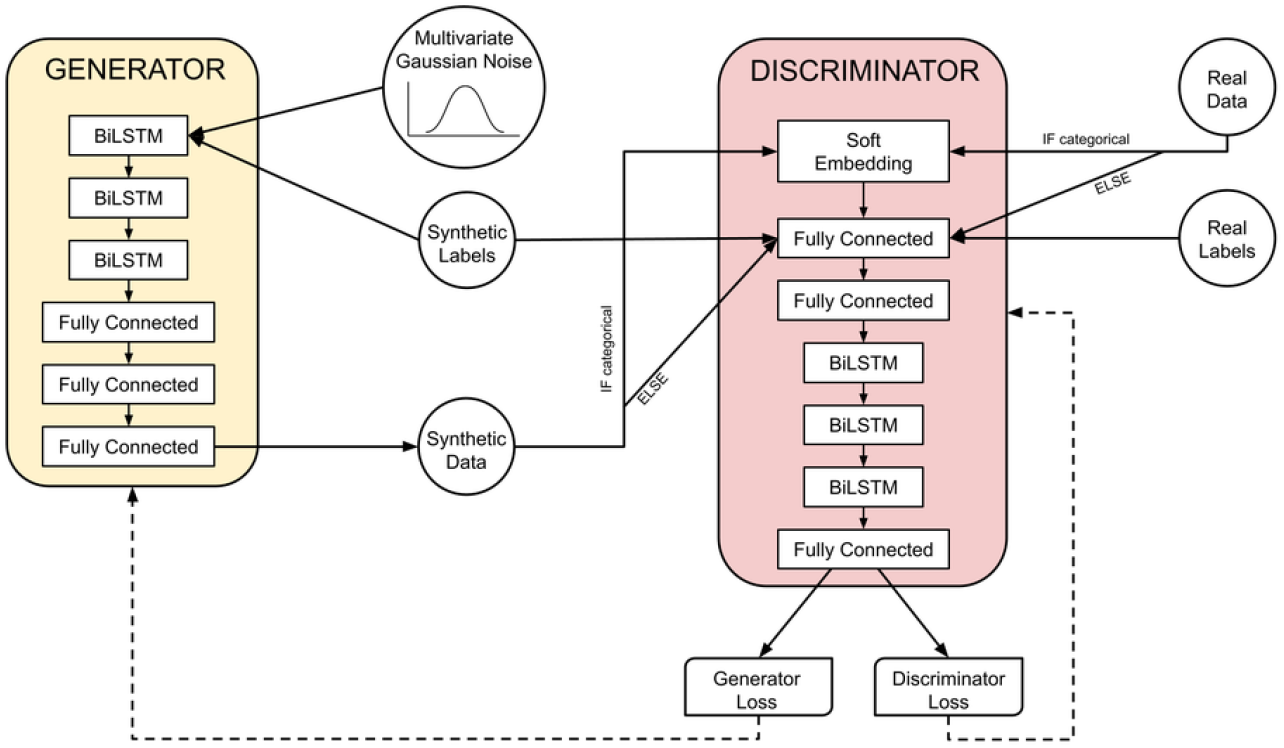
Proposed architecture of our CA-GAN. The Generator and the Discriminator are two deep networks with similar structure and number of parameters. Both employ three stacked Bidirectional LSTMs (BILSTMs) to capture the temporal relationships of longitudinal data. They are trained together adversarially, with a minimax game. Conditioning is achieved with static labels, passed as input to both networks. The Generator also takes Gaussian noise as input and generates time-series data (synthetic patients). The discriminator evaluates the plausibility of the output of the Generator, compared with the real data.

## Data Availability

The data underlying this article are freely available in the MIMIC-III repository

https://mimic.physionet.org/

## Declarations

### 4.8 Ethics approval

The data in MIMIC-III was previously de-identified, and the institutional review boards of the Massachusetts Institute of Technology (No. 0403000206) and Beth Israel Deaconess Medical Center (2001-P-001699/14) both approved the use of the database for research.

### 4.9 Availability of data and materials

The data underlying this article are publicly available in the MIMIC-III repository (https://mimic.physionet.org/).

### 4.10 Code availability

The source code will be made available upon publication at the github repository: https://github.com/nic-olo/CA-GAN

## Supporting information

***Appendix A***

**Distribution Plots for Sepsis**

***Appendix B***

**Datasets**

***Appendix C***

**UMAP and t-SNE parameters**

***Appendix D***

**Joint distributions of variables**

***Appendix E***

**Summary of distance metrics**

***Appendix F***

**Description of Downstream Regression Task**

***Appendix G***

**UMAP Plots**

***Appendix H***

**Absolute Differences in Correlations**

