## Appendices for "Generative AI Mitigates Representation Bias and Improves Model Fairness Through Synthetic Health Data"

### Appendix A Distribution Plots for Sepsis

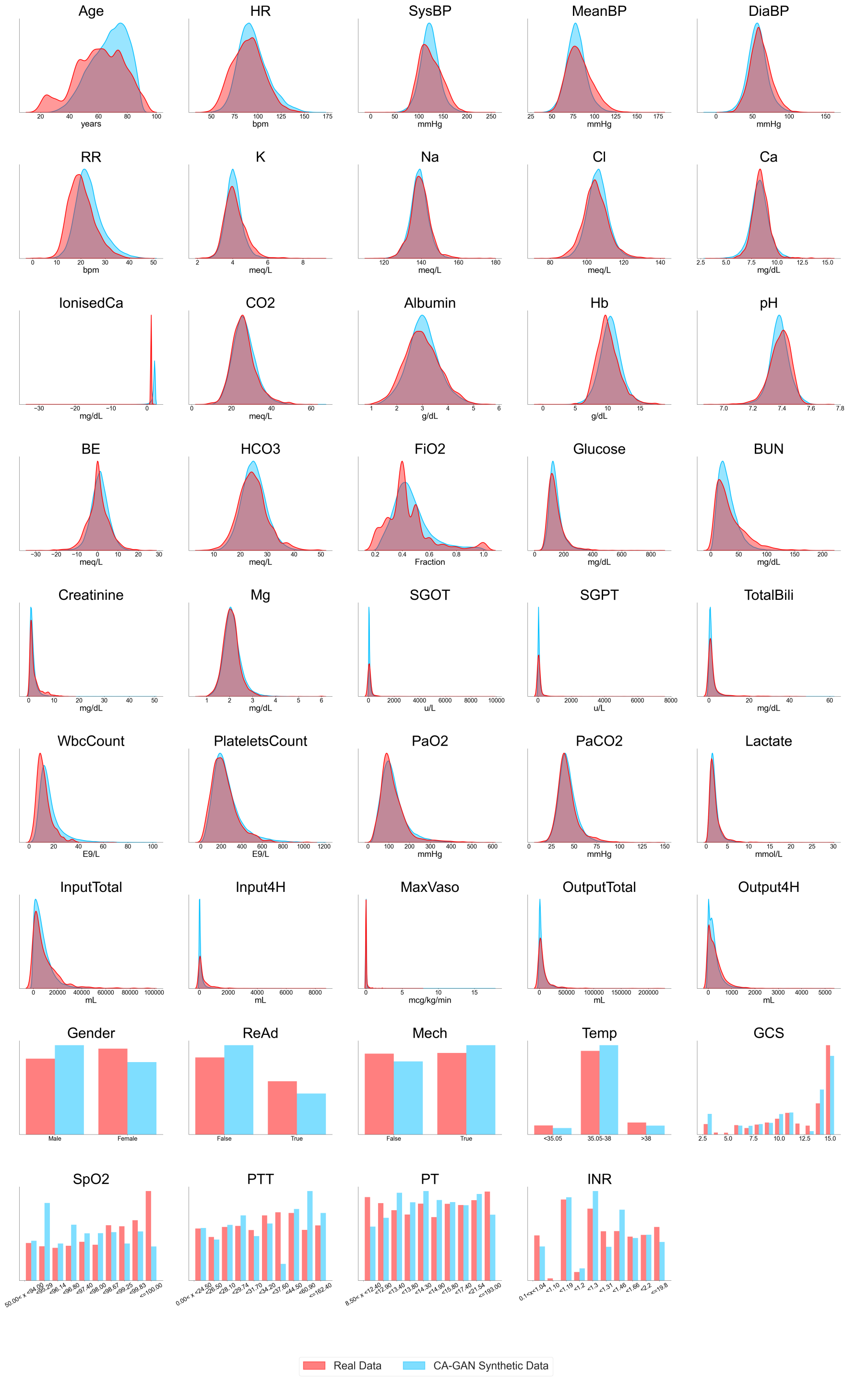


**Fig. A1**: Overlaid distribution plots of real data and CA-GAN synthetic data for each variable in the sepsis dataset.

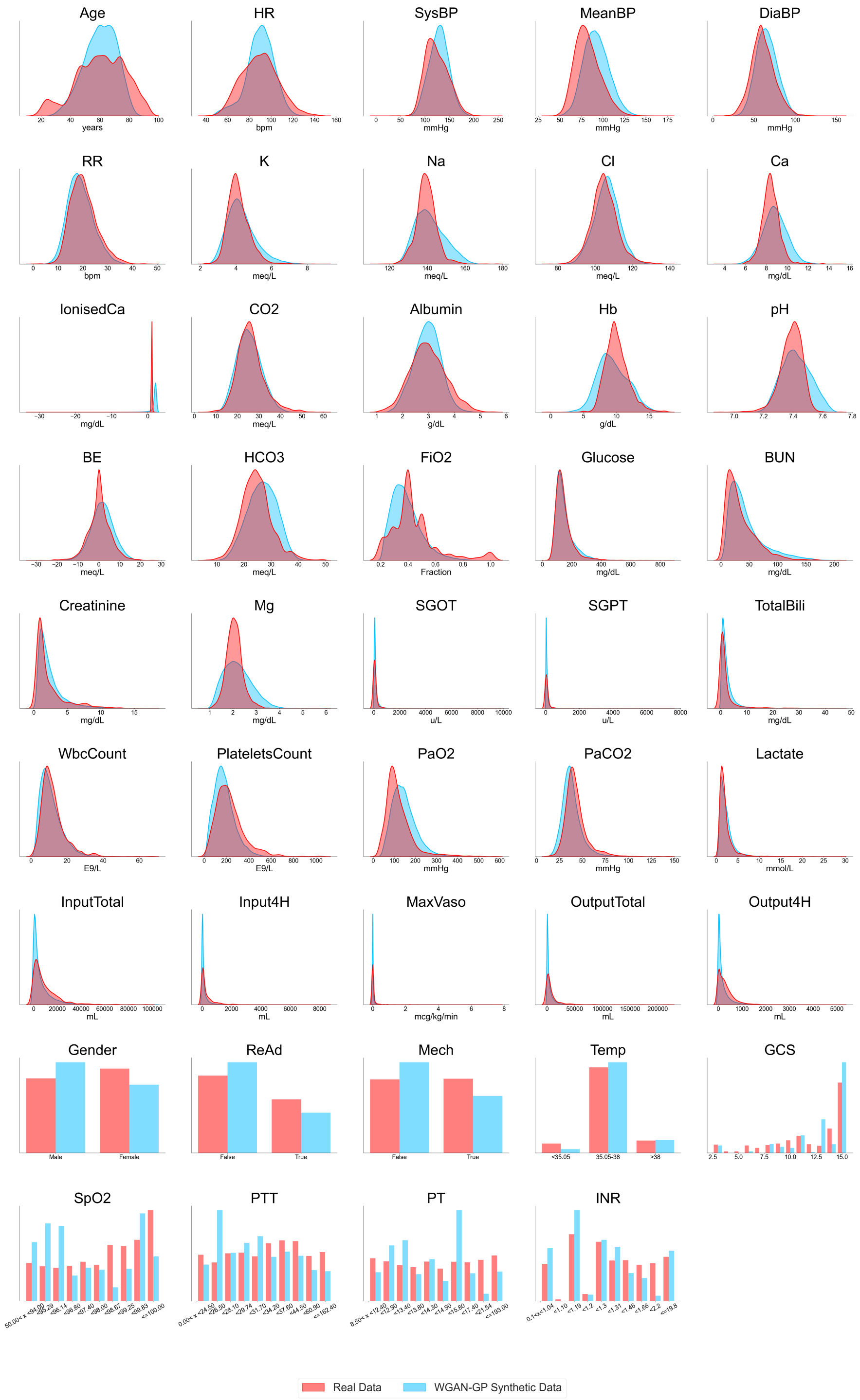


**Fig. A2**: Overlaid distribution plots of real data and WGAN-GP* synthetic data for each variable in the sepsis dataset.

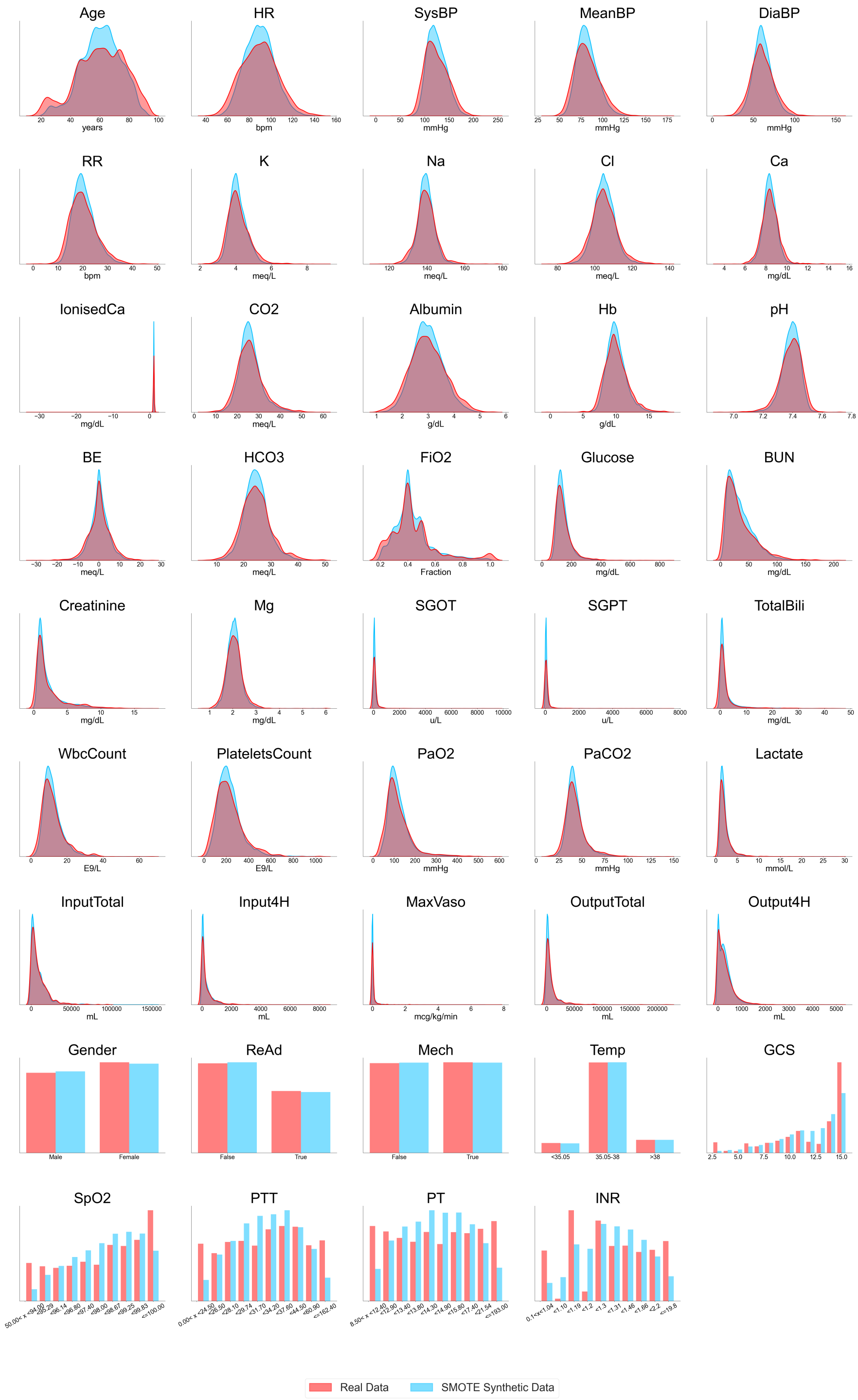


**Fig. A3**: Overlaid distribution plots of real data and SMOTE synthetic data for each variable in the sepsis dataset.

### Appendix B Datasets

**Table B1**: Variables in the acute hypotension dataset. For each variable, the data type, the unit in which it is expressed, and the distribution statistics are presented.

| Variable Name | Data Type | Unit | Descriptive Statistics | |
|---|---|---|---|---|
| Mean Arterial Pressure | numeric | mmHg | Median: 68.00 (Q1: 61.00, Q3: 76.00) | |
| Diastolic Blood Pressure | numeric | mmHg | Median: 54.00 (Q1: 47.00, Q3: 62.00) | |
| Systolic BP | numeric | mmHg | Median: 107.00 (Q1: 96.00, Q3: 120.00) | |
| Urine | numeric | mL | Median: 75.00 (Q1: 45.00, Q3: 140.00) | |
| Alanine Aminotransferase | numeric | IU/L | Median: 35.00 (Q1: 23.00, Q3: 35.00) | |
| Aspartate Aminotransferase | numeric | IU/L | Median: 50.00 (Q1: 33.00, Q3: 50.00) | |
| Partial Pressure of Oxygen | numeric | mmHg | Median: 102.00 (Q1: 99.00, Q3: 102.00) | |
| Lactate | numeric | mmol/L | Median: 1.80 (Q1: 1.20, Q3: 1.80) | |
| Serum Creatinine | numeric | mg/dL | Median: 1.10 (Q1: 0.80, Q3: 1.90) | |
| Fluid Boluses | categorical | mL | 4 Classes | |
| | | | [0,250) : 95.75%; | [250,500) : 0.61% |
| | | | [500,1000) : 1.73%; | ≥ 1000 : 1.91% |
| Vasopressors | categorical | mcg/kg/min | 4 Classes | |
| | | | 0 : 81.95%; | (0,8.4) : 9.02% |
| | | | [8.4,20.28) : 4.51%; | ≥ 20.28 : 4.52% |
| Fraction of Inspired Oxygen | categorical | fraction | 10 Classes | |
| | | | ≤ 0.2 : 0.44%; | 0.2 : 0.45% |
| | | | 0.3 : 5.29%; | 0.4 : 15.52% |
| | | | 0.5 : 61.64%; | 0.6 : 4.36% |
| | | | 0.7 : 2.78%; | 0.8 : 1.43% |
| | | | 0.9 : 1.42%; | 1.0 : 6.67% |
| Glasgow Coma Scale Score | categorical | point | 13 Classes | |
| | | | 3 : 4.57% | 4 : 0.72% |
| | | | 5 : 0.44% | 6 : 2.37% |
| | | | 7 : 3.44% | 8 : 4.03% |
| | | | 9 : 3.63% | 10 : 6.61% |
| | | | 11 : 4.48% | 12 : 1.22% |
| | | | 13 : 3.23% | 14 : 11.48% |
| | | | 15 : 53.80% | |
| Urine Data Measured (M) | binary | - | False: 55.64% | True: 44.36% |
| ALT or AST (M) | binary | - | False: 97.73% | True: 2.27% |
| FiO2 (M) | binary | - | False: 88.80% | True: 11.20% |
| GCS (M) | binary | - | False: 78.06% | True: 21.94% |
| PaO2 (M) | binary | - | False: 95.45% | True: 4.55% |
| Lactic Acid (M) | binary | - | False: 95.61% | True: 4.39% |
| Serum Creatinine (M) | binary | - | False: 92.79% | True: 7.21% |

#### B.1 Data preprocessing

Detailed description of the data preprocessing steps are available from our previous publication [1] in Section 1 of the supplementary material [2]. However, for completeness we highlight some of the main approaches. For the acute hypotension dataset we included adult patients (18 or over) in the MIMIC-III dataset with at least 24 hours of data, aggregating 48 hours of clinical variables from patients with seven or more mean arterial pressure (MAP) values of 65 mmHg or less, indicating acute hypotension. Missing values were replaced with the last available data, while an indicator variable was used to denote whether a value was measured or not. For the sepsis dataset we included adult patients only who had any suspicious infections based on history of administering

[1] Kuo, N.IH., Polizzotto, M.N., Finfer, S. et al. The Health Gym: synthetic health-related datasets for the development of reinforcement learning algorithms. Sci Data 9, 693 (2022). https://doi.org/10.1038/s41597-022-01784-7

[2] https://static-content.springer.com/esm/art%3A10.1038%2Fs41597-022-01784-7/MediaObjects/41597_2022_1784_MOESM4_ESM.pdf

**Table B2**: Variables in the sepsis dataset. For each variable, the data type, the unit in which it is expressed, and the distribution statistics are presented.

| **Variable Name** | **Data Type** | **Unit** | **Descriptive Statistics** | |
|---|---|---|---|---|
| Age | numerical | years | Median: 66.95 (Q1: 54.21, Q3: 78.70) | |
| Heart Rate (HR) | numerical | bpm | Median: 86.75 (Q1: 75.40, Q3: 98.86) | |
| Systolic BP | numerical | mmHg | Median: 118.17 (Q1: 105.33, Q3: 134.20) | |
| Mean BP | numerical | mmHg | Median: 77.25 (Q1: 69.12, Q3: 87.00) | |
| Diastolic BP | numerical | mmHg | Median: 56.81 (Q1: 48.60, Q3: 65.67) | |
| Respiratory Rate (RR) | numerical | bpm | Median: 20.00 (Q1: 16.86, Q3: 23.83) | |
| Potassium (K) | numerical | meq/L | Median: 4.00 (Q1: 3.70, Q3: 4.30) | |
| Sodium (Na) | numerical | meq/L | Median: 139.00 (Q1: 136.00, Q3: 142.00) | |
| Chloride (Cl) | numerical | meq/L | Median: 105.00 (Q1: 101.00, Q3: 109.00) | |
| Calcium (Ca) | numerical | mg/dL | Median: 8.30 (Q1: 7.84, Q3: 8.74) | |
| Ionised Ca | numerical | mg/dL | Median: 1.13 (Q1: 1.08, Q3: 1.18) | |
| Carbon Dioxide (CO2) | numerical | meq/L | Median: 26.00 (Q1: 22.75, Q3: 29.00) | |
| Albumin | numerical | g/dL | Median: 2.90 (Q1: 2.50, Q3: 3.40) | |
| Hemoglobin (Hb) | numerical | g/dL | Median: 10.00 (Q1: 9.04, Q3: 11.20) | |
| pH | numerical | - | Median: 7.40 (Q1: 7.35, Q3: 7.44) | |
| Arterial Base Excess | numerical | meq/L | Median: 0.00 (Q1: -2.00, Q3: 3.15) | |
| Bicarbonate (HCO3) | numerical | meq/L | Median: 25.00 (Q1: 22.00, Q3: 28.00) | |
| FiO2 | numerical | fraction | Median: 0.40 (Q1: 0.40, Q3: 0.50) | |
| Glucose | numerical | mg/dL | Median: 129.67 (Q1: 109.00, Q3: 157.60) | |
| Blood Urea Nitrogen | numerical | mg/dL | Median: 24.00 (Q1: 15.00, Q3: 41.00) | |
| Creatinine | numerical | mg/dL | Median: 1.00 (Q1: 0.70, Q3: 1.60) | |
| Magnesium (Mg) | numerical | mg/dL | Median: 2.03 (Q1: 1.90, Q3: 2.27) | |
| SGOT | numerical | u/L | Median: 41.00 (Q1: 25.00, Q3: 86.00) | |
| SGPT | numerical | u/L | Median: 32.00 (Q1: 18.00, Q3: 69.00) | |
| Total Bilirubin | numerical | mg/dL | Median: 0.70 (Q1: 0.40, Q3: 1.80) | |
| White Blood Cell Count | numerical | E9/L | Median: 11.20 (Q1: 8.20, Q3: 15.20) | |
| Platelets Count | numerical | E9/L | Median: 207.85 (Q1: 141.00, Q3: 296.00) | |
| paO2 | numerical | mmHg | Median: 105.25 (Q1: 82.57, Q3: 140.00) | |
| paCO2 | numerical | mmHg | Median: 40.22 (Q1: 35.25, Q3: 46.09) | |
| Lactate | numerical | mmol/L | Median: 1.60 (Q1: 1.10, Q3: 2.30) | |
| Total Input Fluids | numerical | mL | Median: 6569.08 (Q1: 2540.00, Q3: 13047.66) | |
| Input 4H | numerical | mL | Median: 80.01 (Q1: 20.34, Q3: 327.50) | |
| Max Vasopressors in 4H | numerical | mcg/kg/min | Median: 0.0002 (Q1: 0.00, Q3: 0.0017) | |
| Total Volume Output | numerical | mL | Median: 3893.00 (Q1: 1300.00, Q3: 10070.00) | |
| Output 4H | numerical | mL | Median: 248.00 (Q1: 105.00, Q3: 460.00) | |
| Gender | binary | - | Male: 56.46% | Female: 43.54% |
| Readmission of Patient | binary | - | False: 67.75% | True: 32.25% |
| Mechanical Ventilation | binary | - | False: 46.89% | True: 53.11% |
| Temperature (Temp) | categorical | Celsius | 3 Classes | |
| | | | <35.05: 8.64% | |
| | | | 35.05-38: 79.65% | |
| | | | >38: 11.71% | |
| GCS | categorical | point | 13 Classes | |
| | | | 3: 5.10% | 4: 0.76% |
| | | | 5: 0.93% | 6: 4.51% |
| | | | 7: 3.72% | 8: 4.12% |
| | | | 9: 5.17% | 10: 7.93% |
| | | | 11: 9.68% | 12: 3.98% |
| | | | 13: 3.45% | 14: 13.03% |
| | | | 15: 37.61% | |
| Pulse Oximetry Saturation | categorical | % | 10 Classes | |
| | | | [50.00,94.00): 9.34% | [94.00,95.29): 10.54% |
| | | | [95.29,96.14): 10.11% | [96.14,96.80): 9.49% |
| | | | [96.80,97.40): 9.77% | [97.40,98.00): 8.70% |
| | | | [98.00,98.67): 12.05% | [98.67,99.25): 9.96% |
| | | | [99.25,99.83): 9.99% | [99.83,100.00]: 10.05% |
| Partial Thromboplastin Time | categorical | s | 10 Classes | |
| | | | [0.00,24.50): 9.69% | [24.50,26.50): 9.92% |
| | | | [26.50,28.10): 10.35% | [28.10,29.74): 10.02% |
| | | | [29.74,31.70): 9.76% | [31.70,34.20): 10.09% |
| | | | [34.20,37.60): 10.08% | [37.60,44.50): 10.04% |
| | | | [44.50,60.90): 10.04% | [60.90,162.40]: 9.99% |
| Prothrombin Time | categorical | s | 10 Classes | |
| | | | [8.50,12.04): 10.00% | [12.04,12.90): 9.77% |
| | | | [12.90,13.40): 9.68% | [13.40,13.80): 9.67% |
| | | | [13.80,14.30): 10.54% | [14.30,14.90): 10.03% |
| | | | [14.90,15.80): 10.14% | [15.80,17.40): 9.99% |
| | | | [17.40,21.54): 10.20% | [21.54,193]: 9.98% |
| INR | categorical | - | 10 Classes | |
| | | | [0.10,1.04): 10.00% | [1.04,1.10): 0.48% |
| | | | [1.10,1.19): 17.55% | [1.19,1.20): 2.15% |
| | | | [1.20,1.30): 16.88% | [1.30,1.31): 11.23% |
| | | | [1.31,1.46): 11.68% | [1.46,1.66): 10.02% |
| | | | [1.66,2.2): 9.79% | [2.2,19.8]: 10.22% |

antibiotics. We included all the variables from at least 44 hours before the suspected infection and up to 28 hours after. The missing data was imputed using nearest neighbour method. More detailed information is available in https:

//static-content.springer.com/esm/art%3A10.1038%2Fs41597-022-01784-7/MediaObjects/41597_2022_1784_MOESM4_ESM.pdf

### Appendix C UMAP and t-SNE parameters

In this study, we used t-SNE and UMAP algorithms to perform dimensionality reduction on our datasets and highlight the differences between the results of the three methods under analysis. The following parameters were used for each algorithm:

t-SNE:

Library: scikit-learn version 1.2.2
Parameters for sepsis: $n\_components = 2$, $n\_iter = 500$, $learning\_rate = 100$, $perplexity = 50$
Parameters for acute hypotension: $n\_components = 2$, $n\_iter = 100$, $learning\_rate = 1000$, $perplexity = 30$

UMAP:

Library: umap-learn version 0.5.3
Parameters: $n\_neighbors = 5$, $spread = 5$, $min\_dist = 0.5$

### Appendix D Joint distributions of variables

We have carried out an analysis of a set of variables that are clinically known to follow a joint distribution, namely systolic, diastolic and mean arterial blood pressure. This was to investigate whether CA-GAN can capture joint distributions of variables and whether synthetic data are clinically meaningful, where we know that systolic blood pressure is always higher than diastolic blood pressure. Using a scatter plot in Figure D4 we show that the joint distribution of real data is similar to that of synthetic data.

Following from this, we have also implemented several sanity checks based on clinical knowledge to ensure that the generated synthetic data is clinically meaningful. In this respect, we have performed checks to investigate whether systolic BP values are always lower than diastolic. From our analysis, in the real sepsis dataset, 99.94% of values of these variables were correct; that is, systolic values were always lower than the diastolic values. In the synthetic sepsis dataset, this figure was 99.93%, with only 0.01% difference between the real and the synthetic dataset. On the other hand for the hypotension dataset there were 99.86% correct values of systolic and diastolic variables versus 99.84% in the synthetic dataset, representing a 0.02% difference. This analysis suggests that our architecture captures the structure of the real data quite well.

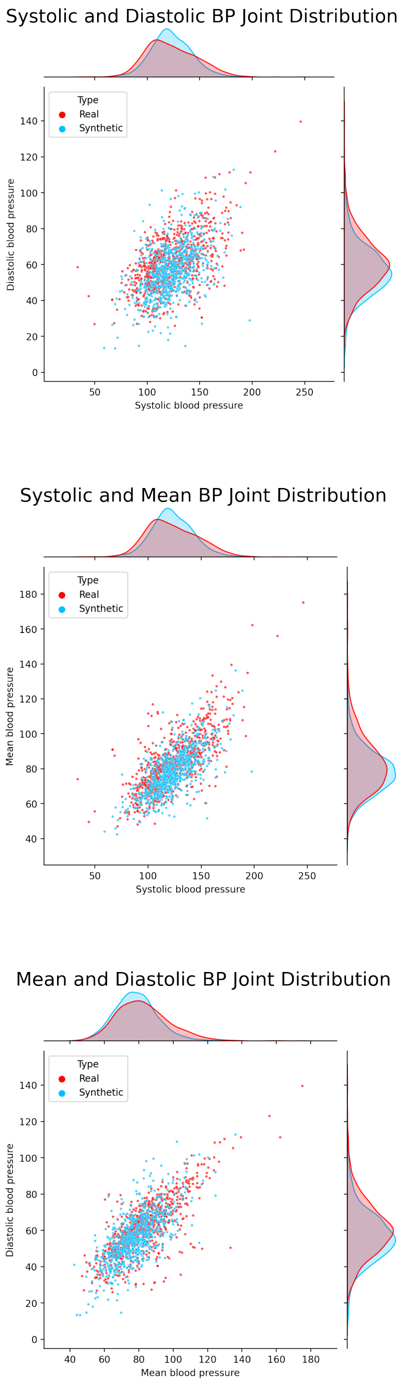


**Fig. D4**: Joint distribution plot of real sepsis data and CA-GAN synthetic data for the variables *Systolic blood pressure*, *Diastolic blood pressure* and *Mean arterial pressure*.

### Appendix E Summary of distance metrics

**Table E3**: Statistics for KL-Divergence and Maximum Mean Discrepancy between the distribution of real and synthetic data.

| | KL-divergence | | | MMD | | |
|---|---|---|---|---|---|---|
| | SMOTE | WGAN-GP* | **CA-GAN** | SMOTE | WGAN-GP* | **CA-GAN** |
| MEDIAN | 0.03407 | 0.02711 | 0.00629 | 0.00752 | 0.00217 | 0.00089 |
| MEAN | 0.05754 | 0.13518 | 0.14128 | 0.01500 | 0.05861 | 0.04790 |
| STD DEV | 0.07222 | 0.22330 | 0.31578 | 0.02385 | 0.09311 | 0.08457 |
| MAX | 0.28191 | 0.91622 | 1.36841 | 0.09954 | 0.25091 | 0.24806 |
| MIN | 0.00068 | 0.00010 | 0.00005 | 0.00047 | 0.00001 | 0.00001 |

(a) Acute hypotension data

| | KL-divergence | | | MMD | | |
|---|---|---|---|---|---|---|
| | SMOTE | WGAN-GP* | **CA-GAN** | SMOTE | WGAN-GP* | **CA-GAN** |
| MEDIAN | 0.02296 | 0.06105 | 0.02292 | 0.00349 | 0.01299 | 0.00377 |
| MEAN | 0.09946 | 0.09282 | 0.06656 | 0.00583 | 0.03848 | 0.02505 |
| STD DEV | 0.29030 | 0.09704 | 0.08906 | 0.00702 | 0.13482 | 0.11603 |
| MAX | 1.88754 | 0.33564 | 0.39555 | 0.02948 | 0.90096 | 0.77536 |
| MIN | 0.00035 | 0.00240 | 0.00199 | 0.00002 | 0.00041 | 0.00003 |

(b) Sepsis data

### Appendix F Description of Downstream Regression Task

For this task, the BiLSTM is trained on the first 20 hours for hypotension and 10 hours for sepsis of the patients' values to predict the next hour, using a sliding window approach. To ensure the fairness of our result, 15% of the time series data points of Black patients and a proportional representation of different genders were set apart as a test set. To account for the stochasticity of the models, we generated five synthetic datasets with CA-GAN. Then, we trained 5 BiLSTM models on each dataset for a total of 25 BiLSTM models (5x(5 per dataset)). The predictions of the trained models were then compared with the test dataset, and the resulting error was averaged across the five models and, subsequently, the five datasets.

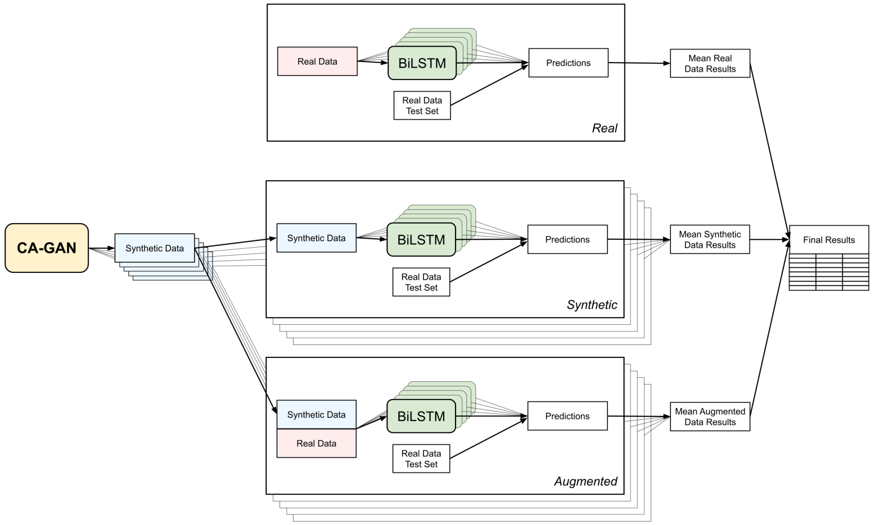


**Fig. F5**: Diagram of BiLSTM Model Training for Continuous Variable Prediction.

### Appendix G UMAP Plots

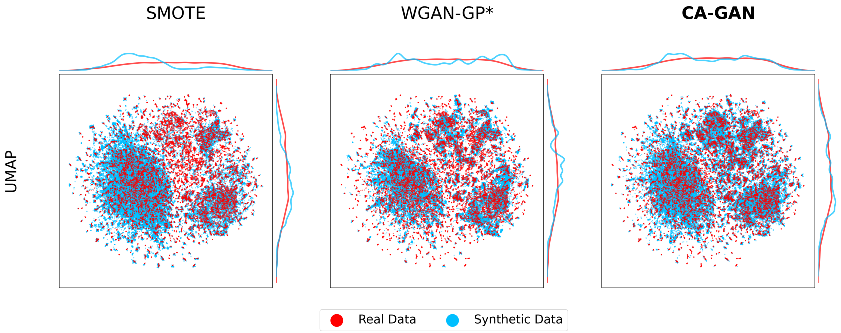


**Fig. G6**: UMAP two-dimensional representations of the acute hypotension dataset for Black patients.

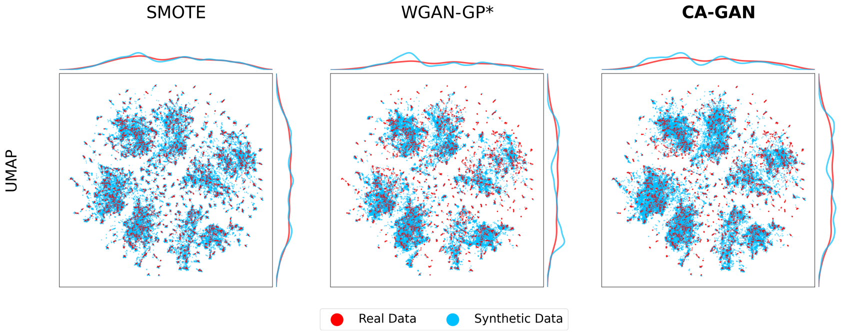


**Fig. G7**: UMAP two-dimensional representations of the sepsis dataset for Black patients.

### Appendix H Absolute Differences in Correlations

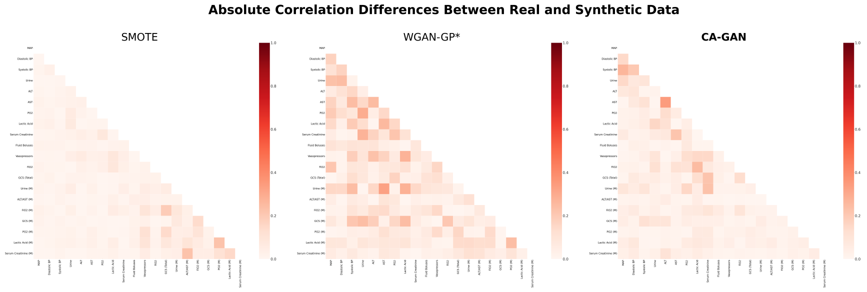


(a) Acute hypotension data

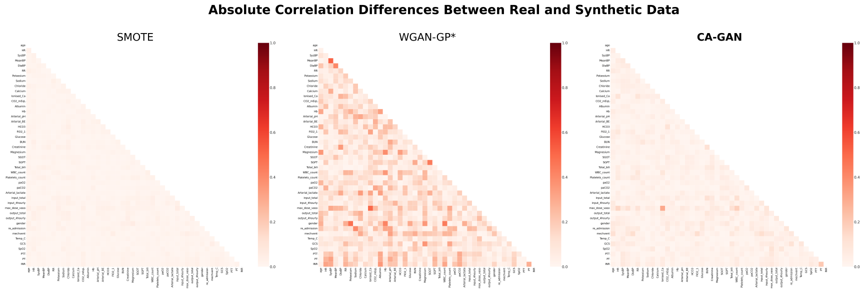


(b) Sepsis data

**Fig. H8**: Absolute difference in correlations between real data and the synthetic data generated with SMOTE, WGAN-GP*, and CA-GAN.
